# Autoantibodies neutralizing type I IFNs in 40% of patients with WNV encephalitis in seven new cohorts

**DOI:** 10.1101/2025.08.31.25334556

**Authors:** Adrian Gervais, Francesca Trespidi, Alessandro Ferrari, Francesca Rovida, Astrid Marchal, Stefania Croce, Irene Cassaniti, Mattia Moratti, Jennifer L. Uhrlaub, David M. Florian, Karin Stiasny, Elisa Burdino, Micol Angelini, Lucy Bizien, Daniele Lilleri, Veronica Codullo, Tal Freund, Yael Paran, Avi Gadoth, Roni Biran, Alessandro Mancon, Camilla Lucca, Stefania Vogiatzis, Monia Pacenti, Mélodie Aubart, Marco Zecca, Patrizia Comoli, Maria Antonietta Avanzini, Jacques Fellay, Antonio Piralla, Francesca Conti, Alberto Dolci, Luisa Barzon, Valeria Ghisetti, Tiziana Lazzarotto, Danilo Cereda, Alessandro Aiuti, Emmanuelle Jouanguy, Paul Bastard, Margaret R. MacDonald, Charles M. Rice, Anne Puel, Laurent Abel, Giada Rossini, Davide Mileto, Yannick Simonin, Anna Nagy, David Hagin, Kristy O. Murray, Fausto Baldanti, Judith H. Aberle, Aurélie Cobat, Shen-Ying Zhang, Jean-Laurent Casanova, Alessandro Borghesi

## Abstract

Mosquito-borne West Nile virus (WNV) infection is a growing global health problem. About 0.5% of infected individuals develop encephalitis. We previously showed that 40% of patients in six cohorts had WNV encephalitis because of circulating auto-antibodies (auto-Abs) neutralizing type I IFNs. In seven new cohorts, we found that the prevalence of auto-Abs was highest (40% [17-44%]) in patients with encephalitis, and very low in a small sample of individuals with asymptomatic or mild infection. In the 13 European, Middle-Eastern and American cohorts available, odds ratios for WNV encephalitis in individuals with these auto-Abs relative to those without them in a large sample of the general population untested for WNV infection range from ∼20 (OR=17.7; 95% CI: 13.8-22.8, *p*<10^−16^) for auto-Abs neutralizing only 100 pg/mL IFN-α2 and/or IFN-ω to >2000 (OR=2218.4; 95% CI: 125.1-39337.7, *p*<10^−16^) for auto-Abs neutralizing high concentrations of IFN-α2 and high or low concentrations of IFN-ω. Pre-existing autoantibodies neutralizing type I IFNs are therefore causal for WNV encephalitis in about 40% of patients.

**Summary:** In 13 cohorts of individuals with WNV infection, the risk of WNV encephalitis is increased 20 to >2,000 times by circulating auto-Abs neutralizing type I IFNs, depending on the concentration and combination of type I IFNs neutralized and patient age.

## Introduction

Mosquito-borne West Nile virus (WNV) is an emerging orthoflavivirus of considerable public health concern, due to its dissemination and its role as a major etiology of epidemic encephalitis worldwide (1,2). In recent years, a wide geographic circulation of WNV across all continents has been observed (3,4,5,6). In 2023, WNV was detected for the first time in 22 new areas of the European Union (EU), corresponding to a 31% increase in the number of affected regions relative to 2022 (1,7). The 2024 transmission season was marked by the broadest ever geographic distribution of WNV in the EU within a single year, with 212 affected regions across 19 countries, versus only 137 regions in 2023 and 173 regions in 2018 (8). Between June and October 2024, the largest outbreak in more than two decades occurred in Israel, with over 930 reported cases and 73 deaths (9). WNV infections have an enormous impact on healthcare systems, with more than 90% of reported cases resulting in hospitalization, neurological manifestations occurring in more than two thirds of cases and a case fatality rate of ∼10% (up to 20% for encephalitis) (8). Tens of thousands of hospitalizations due to WNV infection and thousands of associated deaths have been reported over the last decade in the EU and neighboring countries, in the United States of America (USA), and elsewhere around the world (10,11,12). Seroepidemiological studies have shown that reported cases account for only a small proportion of the WNV infections. Indeed, progression to life-threatening WNV disease (WNVD) requiring hospitalization, particularly encephalitis, occurs in less than 1% of individuals infected with WNV (13). More than 99% of infections are silent or associated with flu-like, self-limiting disease (WNV fever, WNVF) (5,14,15,16,17,18).

The root cause of life-threatening WNVD has long remained elusive (19). We recently showed, in six cohorts from the EU and USA, that blood and CSF autoantibodies (auto-Abs) neutralizing IFN-α2, IFN-β, and/or IFN-ω underlie life-threatening WNVD in ∼35% of hospitalized cases (range: 15-50%) (20). This proportion was higher for patients with WNV neuroinvasive disease, ∼40% of whom (range: 15-55%) were found to have auto-Abs. Auto-Abs neutralizing type I IFNs block the protective antiviral functions of IFN-α2 and/or IFN-β and/or IFN-ω in Vero E6 cells or ARPE-19 cells infected with WNV *in vitro* (21) and in mice *in vivo* (22). They are present in the general population, with a prevalence of <1% in the under-65s and >4% in the over-70s (23). We have shown that they are not induced by WNV infection, and that they precede the appearance of WNV-specific IgM and/or IgG and confer a ∼20- to ∼600-fold increase in the risk of life-threatening WNVD and neuroinvasive disease, depending on the number and concentration of type I IFNs neutralized. These findings, replicated in small case reports (24,25), suggest that auto-Abs neutralizing type I IFNs are strong determinants of WNVD. Moreover, these auto-Abs have been found to underlie tick-borne encephalitis (TBE) in 10% of the most severe cases (26), the most severe cases of the rarer Powassan virus (POWV), Usutu virus (USUV) and Ross River virus (RRV) diseases (27), and ∼35% of cases of life-threatening adverse reactions to yellow fever live-attenuated vaccine-17D strain (YFV-17D) (28). Thus, auto-Abs neutralizing type I IFNs are emerging as strong, common, global determinants of a growing range of arboviral diseases, whether caused by orthoflaviviruses or alphaviruses. We investigated seven new cohorts of individuals infected with WNV, including six recently recruited cohorts established during the seasonal outbreaks in the summers of 2023 and 2024.

## Results

### Seven new cohorts of individuals with WNV infection

We studied 225 subjects hospitalized for life-threatening WNVD from seven new cohorts enrolled at three centers in Italy during 2023 — Pavia (46 subjects), Milan (13 subjects), and Bologna (11 subjects) —one center in Austria (31 subjects, years 2015-2024), one center in Hungary (68 subjects, during 2024), one center in Israel (32 subjects, during 2024) and one center in Texas, USA (24 subjects, years 2002-2008). Neuroinvasive disease was confirmed in 200/225 (89%) patients — encephalitis (154 cases), meningitis (32 cases), acute flaccid paralysis (7 cases) and unspecified neurological disease (7 cases) — whereas there was no reported clinical evidence of neuroinvasive disease in the remaining 25/225 (11%) hospitalized patients. We also enrolled 61 patients with self-limiting, febrile and/or flu-like illness (WNV fever, WNVF), managed as outpatients in Italy (11 subjects), Austria (27 subjects), Hungary (6 subjects), Israel (2 subjects), and the USA (15 subjects), and 32 individuals with recent asymptomatic or paucisymptomatic WNV infection (WNV-infected controls, WNVIC) diagnosed on the basis of the detection of WNV RNA in a nucleic acid amplification test (NAAT) on blood performed at the time of blood donation (Fig. 1A and Fig. S1A-B). For all the individuals enrolled, WNV infection was demonstrated by the presence of WNV-specific IgM or seroconversion to IgG, WNV neutralization assays (29), and/or RT-PCR on serum, plasma, or cerebrospinal fluid samples.

**Figure 1.**
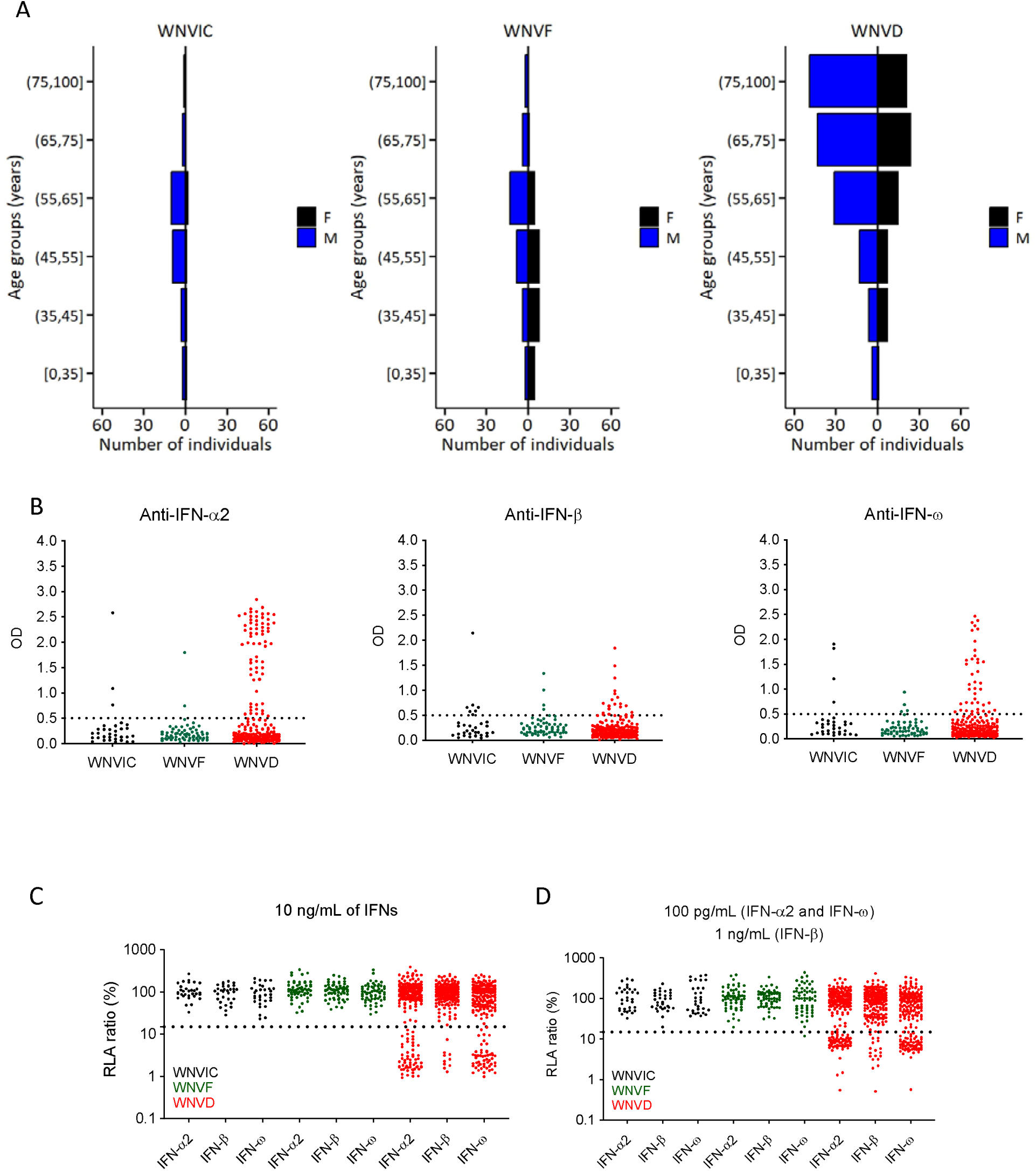
Autoantibodies neutralzing type I IFNs in 318 West Nile virus (WNV)-infected individuals from seven new cohorts. (A) Age and sex distribution of individuals in the WNVIC, WNVF, and WNVD groups. (B) Detection of autoantibodies against IFN-α2, IFN-β, and IFN-ω by ELISA. An OD value > 0.5 (dashed line) indicates a sample considered positive based on the signal typically observed for serum/plasma from healthy donors. (C) Luciferase-based neutralization assay for the detection of autoantibodies neutralizing 10 ng/mL IFN-α2, IFN-ω, or IFN-β. Neutralizing activity was defined as a luciferase signal below 15%. (D) Luciferase-based neutralization assay for detecting autoantibodies neutralizing 100 pg/mL IFN-α2 or IFN-ω, or 1 ng/mL IFN-β. Neutralizing activity was defined as a luciferase signal below 15%.

### Demographics of individuals with WNV infection

The mean age [standard deviation, SD] of patients with WNVD was 68 years [15 years], (range: 10 to 99 years), higher than that for patients with WNVF (51 years [14], range: 19 to 86 years). In the WNVIC group, mean age [SD] was 54 years [11] and age ranged from 32 to 78 years (Fig. S1C). The mean age [SD] of patients with WNVD was higher in the three cohorts enrolled in Italy (Bologna, 77 years [10]), Pavia, 70 years [13] and Milan, 69 years [11], overall: 71 years [13]), than that in the cohorts enrolled in Austria (65 years [13], *p* = 0.04), Hungary (63 years [16], *p* = 8 × 10^−4^) and the USA (62 years [14], *p* = 7 × 10^−3^), but lower than that in the cohort enrolled in Israel (77 years [11], *p* = 0.02) (Fig. S1D). The proportion of male subjects was 65% (147/225) of all patients with WNVD (130/200 [65%] cases of neuroinvasive disease and 102/154 [66%] cases of encephalitis), 54% (33/61) of patients with WNVF, and 84% (27/32) of those with WNVIC (Fig. S1E-F)). Mortality was assessed from vital status data, which were available for 150/200 (75%) patients with neuroinvasive disease. Mortality was 15% (23/150) for the total population of patients with neuroinvasive disease, 17% (19/114) for those with encephalitis, 33% (2/6) for those with acute flaccid paralysis, and 7% (2/27) for those with meningitis; none of the three patients with unspecified neurological disease died and no deaths were reported among patients hospitalized without evidence of neuroinvasive disease (table 1).

**Table 1.** Demographic and clinical characteristics of the study population in the three WNV clinical groups and the two subgroups of WNVD.

| Characteristic | WNVIC<br>*n* = 32<sup>a</sup> | WNVF<br>*n* = 61<sup>a</sup> | WNVD<br>*n* = 225<sup>a</sup> | Subgroups of WNVD |  |
| --- | --- | --- | --- | --- | --- |
|  |  |  |  | Non-neuroinvasive<br>WNVD<br>*n* = 25<sup>a</sup> | Neuroinvasive<br>WNVD<br>*n* = 200<sup>a</sup> |
| **Age (y)** | 54 [11] | 51 [14] | 68 [15] | 68 [12] | 68 [15] |
| **Sex** |  |  |  |  |  |
| F | 5/32 (16%) | 27/61 (44%) | 75/225 (33%) | 8/25 (32%) | 67/200 (34%) |
| M | 27/32 (84%) | 33/61 (54%) | 147/225 (65%) | 17/25 (68%) | 130/200 (65%) |
| Unknown | 0/32 (0%) | 1/61 (2%) | 3/225 (1%) | 0/25 (0%) | 3/200 (1%) |
| **Mortality** | 0/32 (0%) | 0/61 (0%) | 23/225 (10%) | 0/25 (0%) | 23/200 (12%) |
| Unknown | 0/32 (0%) | 0/62 (0%) | 57/225 (25%) | 7/25 (28%) | 50/200 (25%) |
| **Recruitment center** |  |  |  |  |  |
| Italy (PV) | 10/32 (31%) | 5/61 (8%) | 46/225 (20%) | 13/25 (52%) | 33/200 (17%) |
| Italy (MI) | 2/32 (6%) | 6/61 (10%) | 13/225 (6%) | 1/25 (4%) | 12/200 (6%) |
| Italy (BO) | 0/32 (0%) | 0/61 (0%) | 11/225 (5%) | 1/25 (4%) | 10/200 (5%) |
| USA | 0/32 (0%) | 15/61 (25%) | 24/225 (11%) | 0/25 (0%) | 24/200 (12%) |
| Austria | 17/32 (53%) | 27/61 (44%) | 31/225 (14%) | 5/25 (20%) | 26/200 (13%) |
| Israel | 3/32 (9%) | 2/61 (3%) | 32/225 (14%) | 4/25 (16%) | 28/200 (14%) |
| Hungary | 0/32 (0%) | 6/61 (10%) | 68/225 (30%) | 1/25 (4%) | 67/200 (34%) |
| **Year of recruitment** |  |  |  |  |  |
| Before 2018 | 5/32 (16%) | 10/61 (16%) | 27/225 (12%) | 0/25 (0%) | 27/200 (14%) |
| 2018–2022 | 7/32 (22%) | 14/61 (23%) | 11/225 (5%) | 3/25 (12%) | 8/200 (4%) |
| 2023–2024 | 20/32 (63%) | 28/61 (46%) | 187/225 (83%) | 22/25 (88%) | 165/200 (83%) |
| Unknown | 0/32 (0%) | 9/76 (12%) | 0/225 (0%) | 0/25 (0%) | 0/200 (0%) |

WNVIC: West Nile virus-infected controls; WNVF: West Nile virus fever; WNVD: West Nile virus disease PV: Pavia; MI: Milan; BO: Bologna. Mean (SD); or counts (frequency, %).

### Auto-Abs neutralizing type I IFNs are rare in asymptomatic WNVIC

We assessed the prevalence of auto-Abs neutralizing type I IFNs in the three groups of individuals with WNV infection (WNVIC, WNVF and WNVD) from the seven cohorts. We used an enzyme-linked immunosorbent assay (ELISA) to screen serum or plasma samples from all 318 individuals enrolled in this study for circulating IgG auto-Abs against IFN-α2, IFN-β or IFN-ω, with positive results defined as an optical density (OD) > 0.5 (30,31). We then assessed the neutralizing activity of these auto-Abs in a well-established luciferase-based assay (20,23). Blood samples were taken a mean of 18 days (range: 0 to 341 days) after clinical disease onset, or during virological screening before blood donation for WNVIC. In the group of individuals with recent asymptomatic WNV infection (WNVIC), OD values were low for auto-Abs against the three type I IFNs tested in most samples, below the 0.5 OD-unit detection threshold in 25/32 (78%) samples, and slightly above the 0.5 OD-unit detection threshold for at least one type I IFN in 4/32 samples, whereas OD values were very high (> 1.0) for auto-Abs against at least one type I IFN in only 3/32 samples (Fig. 1B). However, neutralizing activity may not be detected in all samples with high (>0.5) or very high (>1.0) OD values on ELISA and may be detected in some with values below the detection threshold (<0.5 OD). We therefore tested 1:10 dilutions of the same serum or plasma samples for the neutralization of high (10 ng/mL) or low (100 pg/mL) concentrations of unglycosylated IFN-α2 and IFN-ω, and high (10 ng/mL) or intermediate (1 ng/mL) concentrations of glycosylated IFN-β. We found no auto-Abs neutralizing high concentrations of IFN-α2 and/or IFN-β and/or IFN-ω or intermediate concentrations of IFN-β in asymptomatic WNVIC, including samples with high OD values (>1.0) on ELISA. At the more physiological concentration of 100 pg/mL, only 1/32 (3%) individuals carried auto-Abs neutralizing IFN-ω only, corresponding to a sample with an OD < 0.5 for auto-Abs against IFN-ω on ELISA (Fig. 1C-D and Fig. 2A-D, table 2). The correlation between ELISA and neutralization results is illustrated in Fig. S2A-C. Auto-Abs neutralizing IFN-α or IFN-ω have recently been shown to block the interaction of the corresponding type I IFNs with both type I IFN receptor subunits (IFNAR1/2), whereas high-titer non-neutralizing auto-Abs limit only the interaction of the corresponding type I IFN with a single receptor subunit (either IFNAR1 or IFNAR2), with the interaction between the IgG and the bound type I IFN being of low avidity (32). This difference may explain the apparent discrepancies between the results obtained by ELISA and those obtained in the neutralization assay for our cohort.

**Figure 2.**
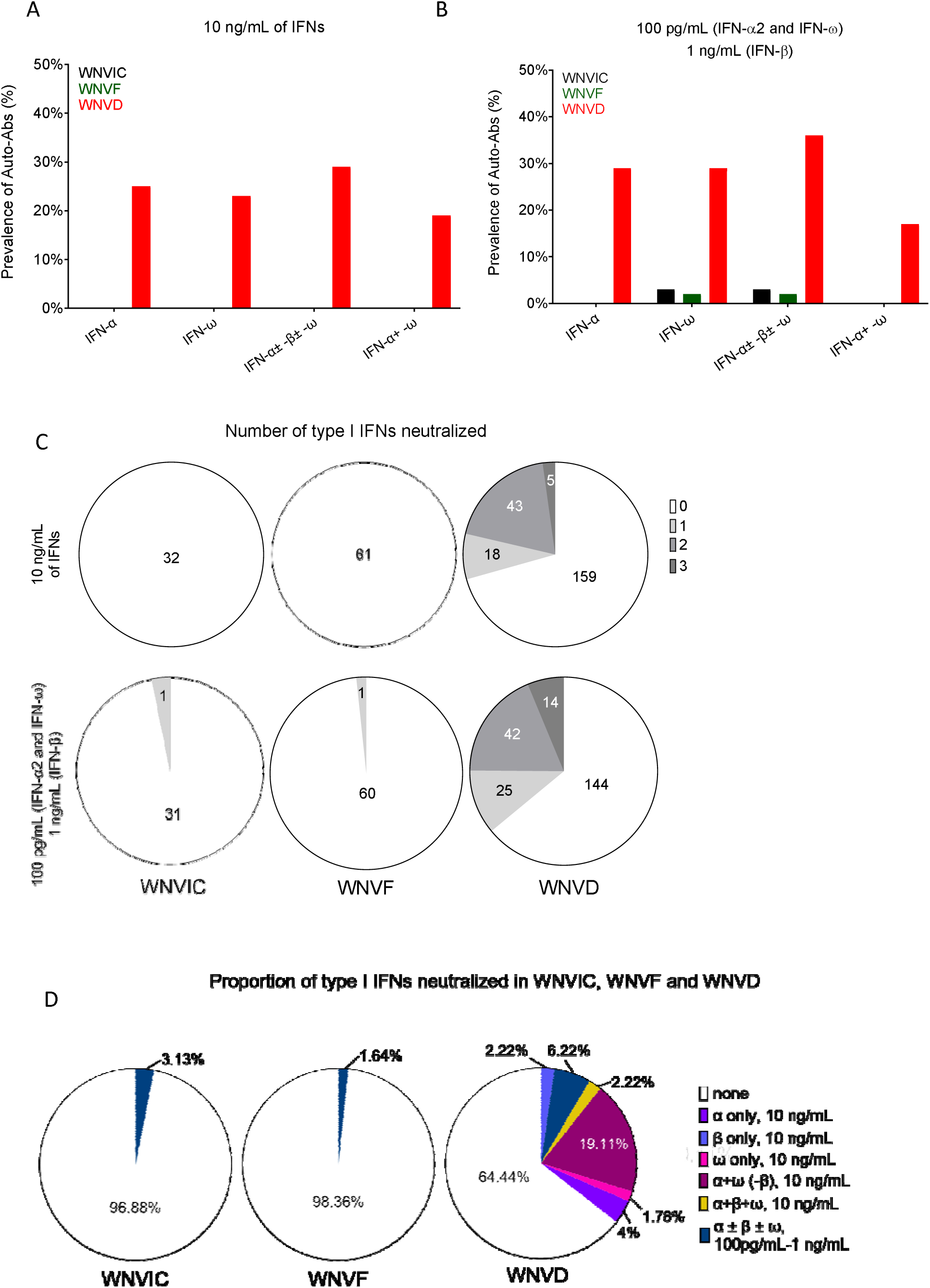
Proportions of individuals with auto-Abs neutralizing type I IFNs in seven new cohorts. (A and B) Frequency of individuals carrying auto-Abs neutralizing type I IFNs at concentrations of 10 ng/mL (A) or 100 pg/mL (B), as determined in luciferase-based neutralization assays, in the three defined groups of WNV-infected individuals: WNVIC, WNVF, and WNVD. IFN-α, auto-Abs neutralizing IFN-α2 (regardless of their effects on other IFNs); IFN-ω, auto-Abs neutralizing IFN-ω (regardless of their effects on other IFNs); IFN-α ± ω ± β, auto-Abs neutralizing IFN-α2 and/or IFN-ω and/or IFN-β; IFN-α + ω, auto-Abs neutralizing both IFN-α2 and IFN-ω. (C) Number of type I IFNs neutralized in the three groups of WNV-infected individuals (WNVIC, WNVF, and WNVD), as determined with the luciferase-based neutralization assay. (D) Proportion of type I IFNs neutralized in the three groups of WNV-infected individuals (WNVIC, WNVF, and WNVD) according to the nature and combination of auto-Abs.

**Table 2.** Type I IFN-neutralizing auto-Abs in the study population.

| Characteristic | Auto-Ab<sup>+</sup>, *n* = 82<sup>a</sup> | No auto-Ab, *n* = 236<sup>a</sup> |
| --- | --- | --- |
| **Age (y)** | 69.52 [15.27] | 60.88 [15.32] |
| ≤40 | 3/29 (10%) | 26/29 (90%) |
| (40–65] | 20/137 (15%) | 117/137 (85%) |
| >65 | 57/148 (39%) | 91/148 (61%) |
| Unknown | 0/4 (0%) | 4/4 (100%) |
| **Sex** |  |  |
| F | 19/107 (18%) | 88/107 (82%) |
| M | 62/207 (30%) | 145/207 (70%) |
| Unknown | 1/4 (25%) | 3/4 (75%) |
| **WNV groups** |  |  |
| WNVD | 81/225 (36%) | 144/225 (64%) |
| WNVF | 1/61 (2%) | 60/61 (98%) |
| WNVIC | 1/32 (3%) | 31/32 (97%) |
| **Subgroups of WNVD** |  |  |
| Non-neuroinvasive WNVD | 9/25 (36%) | 16/25 (64%) |
| **Neuroinvasive WNVD** |  |  |
| All | 71/200 (36%) | 129/200 (64%) |
| WNE | 59/154 (38%) | 95/154 (62%) |
| WNM | 8/32 (25%) | 24/32 (75%) |
| AFP | 2/7 (29%) | 5/7 (71%) |
| UNS | 2/7 (29%) | 5/7 (71%) |

WNVIC: West Nile virus-infected controls; WNVF: West Nile virus fever; WNVD: West Nile virus disease; WNE: WNV encephalitis; WNM: WNV meningitis; AFP: acute flaccid paralysis; UNS: unspecified neurological syndrome Mean (SD), counts or frequency (%)

### Auto-Abs neutralizing type I IFNs are rare in patients with WNVF

Similarly, in the group of patients with mild-to-moderate WNV infection managed as outpatients (WNVF), OD values were low for auto-Abs against the three type I IFNs tested in most samples, below the 0.5 OD detection threshold in 56/61 (92%) samples, and slightly above the 0.5 OD detection threshold for at least one type I IFN in 2/61 (3%) samples, whereas very high OD values (> 1.0) for auto-Abs against at least one type I IFN were obtained for 3/61 (5%) samples (Fig. 1B). The prevalence of samples displaying neutralizing activity against at least one type I IFN remained low in the WNVF group, and similar to that in WNVIC. We detected no auto-Abs neutralizing high concentrations of IFN-α2 and/or IFN-β and/or IFN-ω and/or intermediate concentrations of IFN-β in samples from WNVF patients. We found auto-Abs neutralizing low concentrations of IFN-ω in only 1/61 (2%) samples. The sample testing positive had an OD < 0.5 for anti-IFN-ω IgG auto-Abs on ELISA (Fig. 1C-D and Fig. 2A-D, table 2). The correlation between ELISA and neutralization results is illustrated in Fig. S2A-C. The prevalence of auto-Abs neutralizing type I IFNs was similar in the WNVF and WNVIC groups (1/61 [2%] vs. 1/32 [3%], respectively [*p* = ns]). We detected neutralizing auto-Abs in only 2/93 (2%) individuals with asymptomatic (WNVIC, n = 32) or mild, self-limiting infection (WNVF, *n* = 61) these two groups being analyzed together, a proportion similar to that previously found in the corresponding age groups in the general population (23). Importantly, these two individuals carried auto-Abs neutralizing low concentrations of IFN-ω only, suggesting that auto-Abs neutralizing other type I IFNs — IFN-α2 in particular — could be even rarer in groups of WNV-infected individuals selected on the basis of their resistance to severe disease than in the general population. Thus, the prevalence of auto-Abs neutralizing type I IFNs — typically neutralizing only low concentrations of a single type I IFN (IFN-ω in our sample) — was low in patients with self-limiting WNV infection. These observations also highlight the importance of assessing type I IFN-neutralizing activity (23,33,34) in addition to performing assays detecting the binding of type I IFNs to auto-Abs (e.g., Gyros, ELISA, VIDAS) (30,35,36), when testing samples from individual patients and disease cohorts.

### Auto-Abs neutralizing IFN-**α**2, -**β**, or -**ω** in ∼35% of patients hospitalized for WNVD from seven new cohorts

The prevalence of auto-Abs was much higher in patients who suffered from severe WNVD. In samples from this group of patients, we found OD values above the 0.5 OD-unit detection threshold for IgG auto-Abs against IFN-α2 and/or IFN-β and/or IFN-ω for 73/225 (32%) samples (Fig. 1B and Fig. S3A-B). The prevalence of samples displaying neutralizing activity against at least one type I IFN was high in the WNVD group. We found auto-Abs neutralizing high concentrations of at least one type I IFN in 66/225 (29%) WNVD patients (*p* = 8.45 × 10^−4^ and *p* = 3.29 × 10^−6^ vs. WNVIC and WNVF, respectively), with 57/225 (25%) neutralizing IFN-α2, 52/225 (23%) neutralizing IFN-ω, and 10/225 (4%) neutralizing IFN-β (Fig. 1C-D, Fig. 2A-C and Fig. S4A-D). Specifically, 9/225 (4%) samples neutralized high concentrations of IFN-α2 only, 5/225 (2%) high concentrations of IFN-β only, 4/225 (2%) high concentrations of IFN-ω only, 42/225 (19%) high concentrations of IFN-α2 and IFN-ω, and 6/225 (3%) high concentrations of the three type I IFNs tested; none of the samples neutralized a combination of IFN-α2 and IFN-β only, or a combination of IFN-β and IFN-ω only (Fig. 2D, table 2). Samples with high OD values usually displayed neutralizing activity (Fig. S2A-C, table 2). In addition, 3/42 (7%) patients carrying auto-Abs neutralizing high concentrations of both IFN-α2 and IFN-ω, 1/9 (11%) patients neutralizing high concentrations of IFN-α2 only and 4/159 (3%) patients without auto-Abs neutralizing high concentrations of any of the three type I IFNs tested also displayed neutralization of intermediate but not high concentrations of IFN-β. Overall, samples from 9/225 (4%) patients neutralized intermediate, but not high concentrations of IFN-β, regardless of the neutralization of other type I IFNs. At more physiological concentrations, 81/225 (36%) samples from patients hospitalized for life-threatening WNVD neutralized at least 100 pg/mL IFN-α2 and/or IFN-ω and/or 1 ng/mL IFN-β, including 12/225 (5%) samples neutralizing 100 pg/mL of IFN-α2 only, 11/225 samples (4%) neutralizing 100 pg/mL IFN-ω only, 2/225 (<1%) neutralizing 1 ng/mL IFN-β only, 39/225 (17%) neutralizing 100 pg/mL of both IFN-α2 and IFN-ω, 1/225 (<1%) neutralizing both 100 pg/mL IFN-α2 and 1 ng/mL IFN-β, 2/225 (1%) neutralizing both 100 pg/mL IFN-ω and 1 ng/mL IFN-β, and 14/225 (7%) neutralizing 100 pg/mL IFN-α2 and IFN-ω and 1 ng/mL IFN-β. In total, 17/225 (8%) samples neutralized 100 pg/mL IFN-α2 and/or IFN-ω and/or 1 ng/mL IFN-β but did not neutralize high concentrations of any of the three type I IFNs tested.

### Auto-Abs neutralizing IFN-**α**2, -**β**, or -**ω** in ∼40% of patients with WNV neuroinvasive disease

Among WNVD patients, the prevalence of auto-Abs was higher in patients with WNV neuroinvasive disease (WNV encephalitis, meningitis or acute flaccid paralysis), with 64/200 (32%) having an OD>0.5 on ELISA, and 58/200 (29%) displaying the neutralization of high concentrations of at least one of the three type I IFNs tested (*p* = 9.75 × 10^−4^ and *p* = 4.37 × 10^−6^ vs. WNVIC and WNVF, respectively). In this subgroup of patients, samples from 71/200 (36%) patients contained circulating auto-Abs neutralizing at least 100 pg/mL IFN-α2 and/or IFN-ω and/or 1 ng/mL IFN-β. The prevalence of individuals carrying auto-Abs neutralizing type I IFNs was highest in the subgroup of patients suffering from encephalitis — the most severe neuroinvasive manifestation of WNV infection — with 52/154 (34%) samples displaying an OD>0.5 on ELISA. In this subgroup of patients, samples from 49/154 (32%) patients neutralized high concentrations of at least one of the three type I IFNs tested. In tests of the neutralization of more physiological concentrations, samples from 59/154 (38%) patients contained auto-Abs neutralizing at least 100 pg/mL IFN-α2 and/or IFN-ω and/or 1 ng/mL IFN-β, a prevalence higher than that in patients with acute flaccid paralysis (2/7 [29%]), meningitis (8/32 [25%]), or unspecified neurological disease (2/7 [29%]), and hospitalized patients with no documented neuroinvasive disease (9/25 [36%]). Finally, for patients with available mortality data, auto-Abs neutralizing 100 pg/mL IFN-α2 and/or IFN-ω and/or 1 ng/mL of IFN-β were detected in 43/127 (34%) patients with neuroinvasive disease (36/95 [38%] with encephalitis) who survived, and in 10/23 (44%) patients with neuroinvasive disease (8/19 [42%] with encephalitis) who died from WNV infection. Overall, consistent with our previous report (20), we found auto-Abs neutralizing IFN-α2 and/or IFN-β and/or IFN-ω in almost 40% of patients with WNV neuroinvasive disease, including encephalitis in particular, from seven new cohorts enrolled in five countries, including two new geographic areas.

### WNV infection in 981 individuals from nine centers in five countries

We then performed a combined analysis on the six previously reported cohorts (20) and the seven new cohorts consisting of 981 individuals with WNV infection recruited in five countries. There were 666 patients with WNVD (including 548 suffering from neuroinvasive disease, 376 of whom had documented WNV encephalitis), 169 patients managed as outpatients for WNVF, and 146 WNVIC (Fig. S5A-B). Mean age, sex distribution and the prevalence of auto-Abs by disease group were comparable between the previously studied and new cohorts, even after analysis by country or by recruitment center (Fig. S5C-F). We performed subsequent analyses on the whole cohort of 666 WNVD. In total, 423 of these patients were recruited in Italy: 116 in Pavia (20 in 2018, 50 in 2022, 46 in 2023), 196 in Padua (89 in 2018, 107 in 2022), 58 in Bologna (47 in 2022, 11 in 2023), 40 in Turin (29 in 2018, 11 in 2022), and 13 in Milan (13 in 2023), 99 patients were recruited in Budapest, Hungary (20 in 2018, 11 in 2019, 68 in 2024), 32 were recruited in Tel Aviv, Israel in 2024, 31 were recruited in Vienna, Austria, between 2015 and 2024, and 81 were recruited in Houston, Texas, USA, between 2002 and 2010. In total, 84/666 (13%) patients were recruited between 2002 and 2017, 165/666 (25%) were recruited in 2018, 12/666 (18%) were in 2019, 1/666 was recruited in 2021, 217/666 (33%) were recruited in 2022, 71/666 (11%) were recruited in 2023 and 116/666 (17%) were recruited in 2024.

### Auto-Abs neutralizing IFN-**α**2, -**β**, or -**ω** in 981 individuals with WNV infection, by country, center and year of recruitment

We then analyzed the prevalence of auto-Abs neutralizing low concentrations of IFN-α2 and/or IFN-ω and/or intermediate concentrations of IFN-β in WNVD patients by country and year of recruitment. The overall prevalence of auto-Abs neutralizing at least one type I IFN was 236/666 (35%) in WNVD patients. The highest prevalence was found in WNVD patients from Israel (13/32 [40%]) and Italy (165/423 [39%], including 7/13 [54%] in Milan, 54/116 [47%] in Pavia, 17/40 [43%] in Turin, 25/58 [38%] in Bologna, 65/196 [33%] in Padua), followed by Austria (11/31 [35%]), Hungary (32/99 [32%]) and the USA (14/81 [17%]). The overall prevalence of auto-Abs neutralizing at least one type I IFN was 206/548 (38%) in patients with WNV neuroinvasive disease (135/376 [36%] in WNV encephalitis patients), and was highest in Israel (12/28 [43%]) and Italy (141/321 [44%], including 6/12 [50%] in Milan, 47/94 [50%] in Pavia, 16/38 [42%] in Turin, 22/52 [42%] in Bologna, 50/125 [40%] in Padua), followed by Austria (10/26 [38%]), Hungary (29/92 [32%]) and the USA (14/81 [17%]) (Fig. 3A-B). The prevalence of auto-Abs neutralizing at least one type I IFN was 16/84 (19%) in WNVD patients recruited between 2002 and 2017, 67/165 (41%) in those recruited in 2018, 5/12 (42%) in 2019, 1/1 (100%) in 2021, 78/217 (36%) in 2022, 34/71 (48%) in 2023, 35/116 (30%) in 2024 (Fig. 3C-D). The proportion of individuals with WNVF from the new cohorts carrying auto-Abs was similar to that in the general population (1/61 [2%]), and lower than that in WNVF patients from the previously reported cohorts (15/108 [14%]) (20). In the WNVF patients of the previously reported cohorts, substantial differences were observed between recruitment centers (Fig. 3A-F), possibly reflecting differences in the clinical criteria used to define WNVF and disease severity between centers and across years of recruitment, or misclassification due to missing follow-up data and the unreported clinical worsening and hospitalization of some patients initially tested for WNVF.

**Figure 3.**
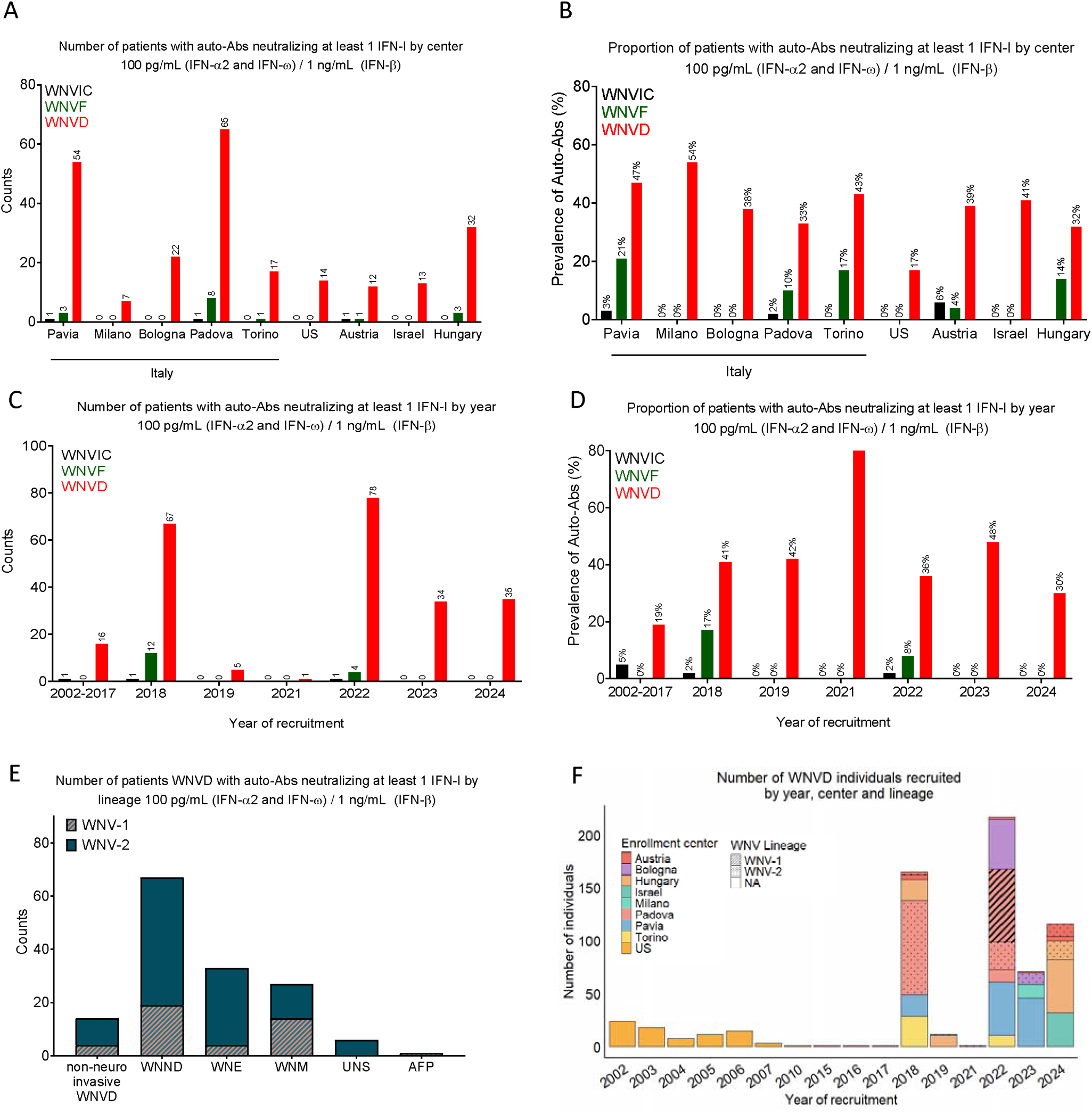
Proportions of patients with auto-Abs neutralizing type I IFNs by center and year of enrollment, and lineage in 13 cohorts from 5 countries. (A-B) Number and prevalence of individuals with auto-Abs neutralizing at least one type I IFN at a concentration of 100 pg/mL (IFN-α2, IFN-ω) or 1 ng/mL (IFN-β) in the three groups of individuals infected with WNV (WNVIC, WNVF, WNVD), by enrollment center. (C-D) Number and prevalence of individuals with auto-Abs neutralizing at least one type I IFN at a concentration of 100 pg/mL (IFN-α2, IFN-ω) or 1 ng/mL (IFN-β) in the three groups of individuals infected with WNV (WNVIC, WNVF, WNVD), by enrollment year. (E) Number of individuals with auto-Abs neutralizing at least one type I IFN at a concentration of 100 pg/mL (IFN-α2, IFN-ω) or 1 ng/mL (IFN-β) in subgroups of WNND patients: WNV encephalitis (WNE), WNV meningitis (WNM), acute flaccid paralysis (AFP) and unspecified neurological syndrome (UNS), by WNV lineage. (F) Distribution by year (*x*-axis), recruitment center (colors), and infecting WNV virus lineage (WNV-1, dashed boxes, WNV-2, dotted boxes) of the individuals with WNVD recruited.

### Auto-Abs neutralizing IFN-**α**2, -**β**, or -**ω**, by infecting WNV lineage

We also analyzed the prevalence of auto-Abs according to the infecting WNV lineage, determined for 66/146 (45%) WNVIC, 84/169 (50%) patients with WNVF and 230/666 (35%) WNVD patients. The proportions of individuals infected with WNV-1 or WNV-2 were comparable in the three WNV infection groups. WNV-1 was detected in 16/66 (24%) WNVIC, in 21/84 (25%) patients with WNVF, and in 69/230 (30%) of the samples obtained from WNVD patients during the 2022 summer outbreak in Padua. WNV-2 was detected in 50/66 (76%) WNVIC, 63/84 (75%) patients with WNVF, and in 161/230 (70%) samples from WNVD patients from Austria, Hungary, Bologna and Padua obtained in 2018, 2019, 2021, 2022, 2023 and 2024 (Fig. 3E-F). We found auto-Abs neutralizing low concentrations of IFN-α2 and/or IFN-ω and/or intermediate concentrations of IFN-β in 23/69 (33%) WNVD patients infected with the WNV-1 lineage, including 19/56 (34%) patients with WNV neuroinvasive disease (14/35 [40%] with meningitis, 1/2 [50%] with acute flaccid paralysis, and 4/19 [21%] with encephalitis), and in 58/161 (36%) WNVD patients infected with the WNV-2 lineage, including 48/104 (46%) patients with WNV neuroinvasive disease (13/31 [42%] with meningitis, none (0/1) with acute flaccid paralysis, 6/19 [32%] with unspecified neurological syndrome, and 29/53 [55%] with encephalitis) (Fig. 3E-F, table S1). Thus, we found no substantial difference in the prevalence of auto-Abs between WNVD patients those infected with WNV-1 and WNVD patients infected with WNV-2 viruses (23/69 [33%] vs 58/161 [37%], *p* = 0.81). The higher prevalence of individuals carrying auto-Abs among those with WNV neuroinvasive disease due to WNV-2 than among those with WNV neuroinvasive disease due to WNV-1 viruses, particularly for patients with encephalitis (29/53 [55%] vs. 4/19 [21%], *p* = 0.02), requires confirmation in future studies and may reflect sample selection at diagnosis for virological studies, or, alternatively, differences in the neurotropism of the viral lineages in individuals with impaired type I IFN immunity in the blood or at the blood-brain barrier due to circulating auto-Abs.

### Higher prevalence of auto-Abs in male individuals

We previously showed that age and sex influence the prevalence of auto-Abs neutralizing type I IFNs. We therefore analyzed the effects of both variables on the prevalence of auto-Abs neutralizing low concentrations of IFN-α2 and/or IFN-ω, and/or intermediate concentrations of IFN-β, in each disease category. For 2/225 WNVD patients, sex and age were not reported, for 2/225 additional WNVD patients sex was reported but not age, and for 1/61 WNVF patients, age was reported but not sex. In the WNVD group, the prevalence of auto-Abs was higher in male subjects than in female subjects in the new cohorts (60/147 [41%] vs. 19/76 [25%], OR = 2.07, 95% CI: 1.13–3.89, *p* = 0.02), and in the total cohort of 664/981 patients with WNVD and for whom sex data were available (172/433 [40%] vs. 63/231 [27%], OR = 1.75, 95% CI: 1.24–2.48, *p* = 1.7 × 10^−^

^3^). A similar trend was observed in the neuroinvasive disease group in the new cohorts (53/130 [41%] male patients vs. 17/68 [25%] female patients, OR = 2.06, 95% CI: 1.09–4.04, *p* = 0.03) and in the total cohort of 546/981 patients with neuroinvasive disease and available sex data (149/362 [41%] male patients vs. 56/184 [30%] female patients, OR = 1.59, 95% CI: 1.09–2.33, *p* = 0.02). By contrast, no difference in the prevalence of the auto-Abs between the sexes was observed in WNVIC (1/27 [4%] male patients vs. 0/5 female patients, *p* = 1.0) or in the WNVF group (1/33 [3%] male patients vs. 0/27 female patients, *p* = 1.0) in the seven new cohorts, and in WNVIC (3/114 [3%] male patients vs. 0/32 female patients, *p* = 1) or the WNVF group (11/88 [13%] male patients vs. 5/80 [6%] female patients, *p* = 0.17) in the total cohort (Fig. 4A, table S2).

**Figure 4.**
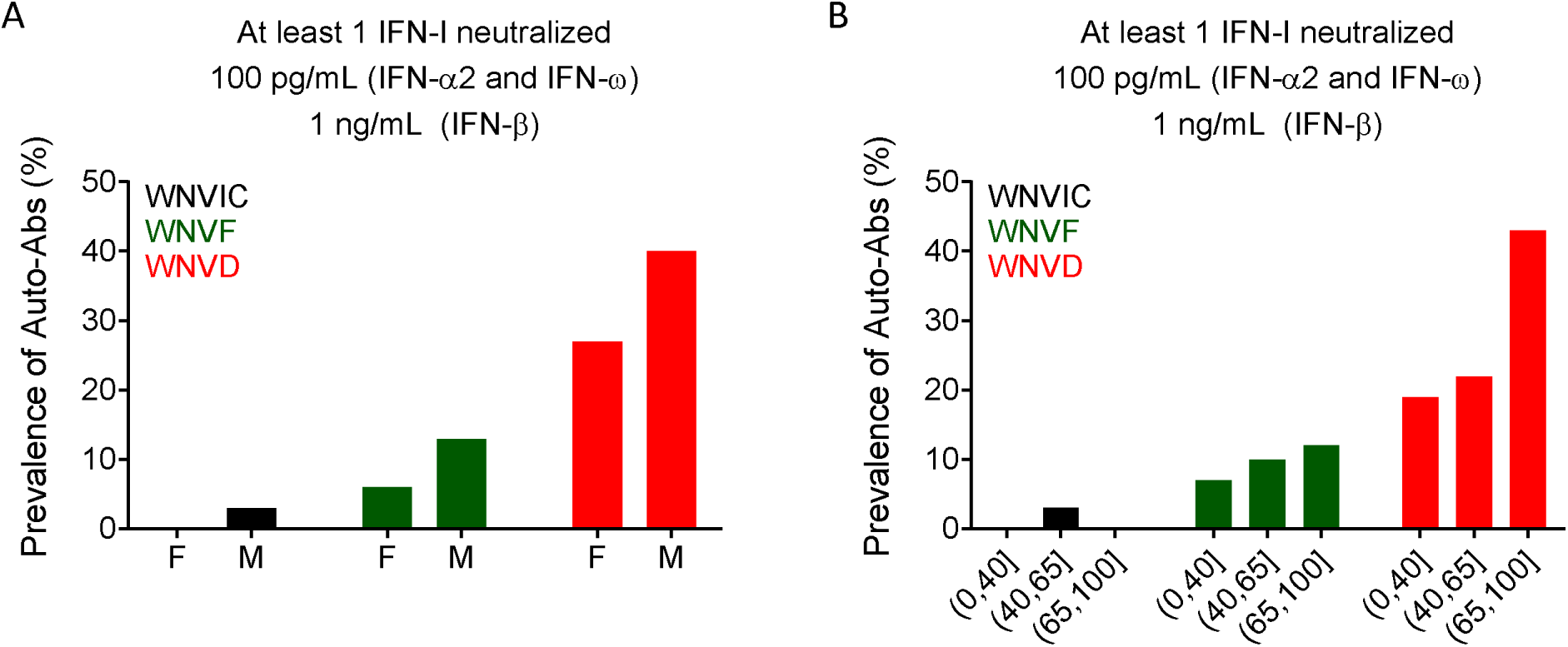
Proportions of patients with auto-Abs neutralizing type I IFNs by sex and age in 13 cohorts from 5 countries. (A) Prevalence of individuals with auto-Abs neutralizing at least one type I IFN at a concentration of 100 pg/mL (IFN-α2, IFN-ω) or 1 ng/mL (IFN-β) in the three groups of individuals infected with WNV (WNVIC, WNVF, WNVD), by sex. (B) Prevalence of individuals with auto-Abs neutralizing at least one type I IFN at a concentration of 100 pg/mL (IFN-α2, IFN-ω) or 1 ng/mL (IFN-β) in the three groups of WNV-infected individuals (WNVIC, WNVF, WNVD), by age class.

### Higher prevalence of auto-Abs in individuals over the age of 65 years

The individuals with neutralizing auto-Abs were significantly older than those without them in the seven new cohorts (mean age [SD], 70 [15] vs. 61 [15] years; *p* = 2.43 × 10^−5^) and in the total cohort of 977/981 individuals for whom age data were available (mean age [SD], 71 [15] vs. 60 [17] years; *p* = 2.18 × 10^−19^). This difference was driven primarily by the older age of the individuals with auto-Abs in the WNVD group (mean age [SD], 72 [14] vs. 65 [16] years; *p* = 1.9 × 10^−7^ in the group of 662 WNVD patients with available age data). By contrast, mean age did not differ significantly between individuals with and without auto-Abs in the group of 168 individuals with WNVF and available age data (mean age [SD], 57 [18] vs. 54 [18] years; *p* = 0.41). Stratification by age group (≤40, (40–65], and >65 y) in the new cohorts confirmed our previous observations [19], with auto-Abs neutralizing low concentrations of IFN-α2 and/or IFN-ω, and/or intermediate concentrations of IFN-β being more frequent in patients with severe disease, particularly older individuals. In individuals aged ≤40 years, no auto-Abs were detected in the WNVIC group, whereas auto-Abs were present in 3/45 (7%) WNVF patients and 8/42 (19%) WNVD patients. The prevalence of auto-Abs increased with age. In the 40–65 years age group, auto-Abs were detected in 3/105 (3%) WNVIC, 8/83 (10%) WNVF patients and 45/203 (22%) WNVD patients. The prevalence of the auto-Abs increased sharply after the age of 65 years, with auto-Abs detected in 0/14 WNVIC, 5/41 (12%) WNVF and 181/417 (43%) WNVD cases (Fig. 4B). Thus, the risk of carrying auto-Abs increased with age, and was higher in subjects >65 y old than in subjects ≤65 y old in the WNVD group (181/417 [43%] vs. 53/245 [22%], OR = 2.78, 95% CI: 1.95–4.01; *p* = 2.77 × 10^−8^), in patients with neuroinvasive disease (162/362 [45%], vs. 42/182 [23%] OR = 2.7, 95% CI: 1.82–4.07; *p* = 1.30 × 10^−6^), and in patients with encephalitis (108/262 [41%] vs. 25/111 [23%], OR = 2.78, 95% CI: 1.95–4.01; *p* = 2.77 × 10^−8^). The prevalence of these auto-Abs was highest in males >65 y of age with encephalitis (81/173 [47%]).

### Auto-Abs neutralizing type I IFNs are as rare in WNVIC and patients with WNVF as in the general population

For estimation of the risk of clinical disease conferred by the presence and nature of auto-Abs neutralizing type I IFNs, we compared the proportions of subjects carrying various types and combinations of neutralizing auto-Abs with the proportions in individuals carrying the corresponding neutralizing auto-Abs among 34,159 healthy men and women aged 18–100 years from the French general population, after adjustment for age and sex (23). We found no significant difference in the prevalence of neutralizing auto-Abs between WNVIC and the general population, regardless of the type or combination of auto-Abs considered, in both a separate analysis of the new cohorts (*n* = 32 WNVIC) and a combined analysis of these and the previously described cohorts (*n* = 146 WNVIC) (1/32 [3%] vs. 295/13570 [2%], OR = 1.97, 95% CI: 0.39-9.94, *p* = 0.46; and 3/146 [2%] vs. 295/13570 [2%], OR = 1.04, 95% CI: 0.36-3.01, *p* = 0.95, for auto-Abs neutralizing 100 pg/mL IFN-α2 and/or IFN-ω for the new cohorts and the total cohort, respectively), consistent with the absence of clinical disease despite documented WNV infection in this group (Figs. 3A-B and Fig. 5). Similarly, we found no significant difference in the prevalence of neutralizing auto-Abs between individuals with WNVF in the new cohorts (*n* = 61 WNVF) and individuals in the general population, regardless of the type or combination of auto-Abs considered (1/61 [2%] vs. 295/13570 [2%], OR = 1.12, 95% CI: 0.22-5.63, *p* = 0.89 for auto-Abs neutralizing 100 pg/mL IFN-α2 and/or IFN-ω) (Fig. 5 and 6C, table 3). This finding contrasts with the higher prevalence of auto-Abs neutralizing various combinations of type I IFNs in patients with WNVF in our previous study (15/108 [14%]) (20), and therefore in the combined analysis of the new and previously reported cohorts (*n* = 169 WNVF), resulting in a ∼5-20-times increase in the risk of WNVF depending on the type and nature of the neutralizing auto-Abs (16/169 [10%] vs. 295/13570 [2%], OR = 4.7, 95% CI: 2.8-7.9, *p* = 1.37 × 10^−6^ for auto-Abs neutralizing 100 pg/mL IFN-α2 and/or IFN-ω), (Fig. 5 and 6D, table 3).

**Figure 5.**
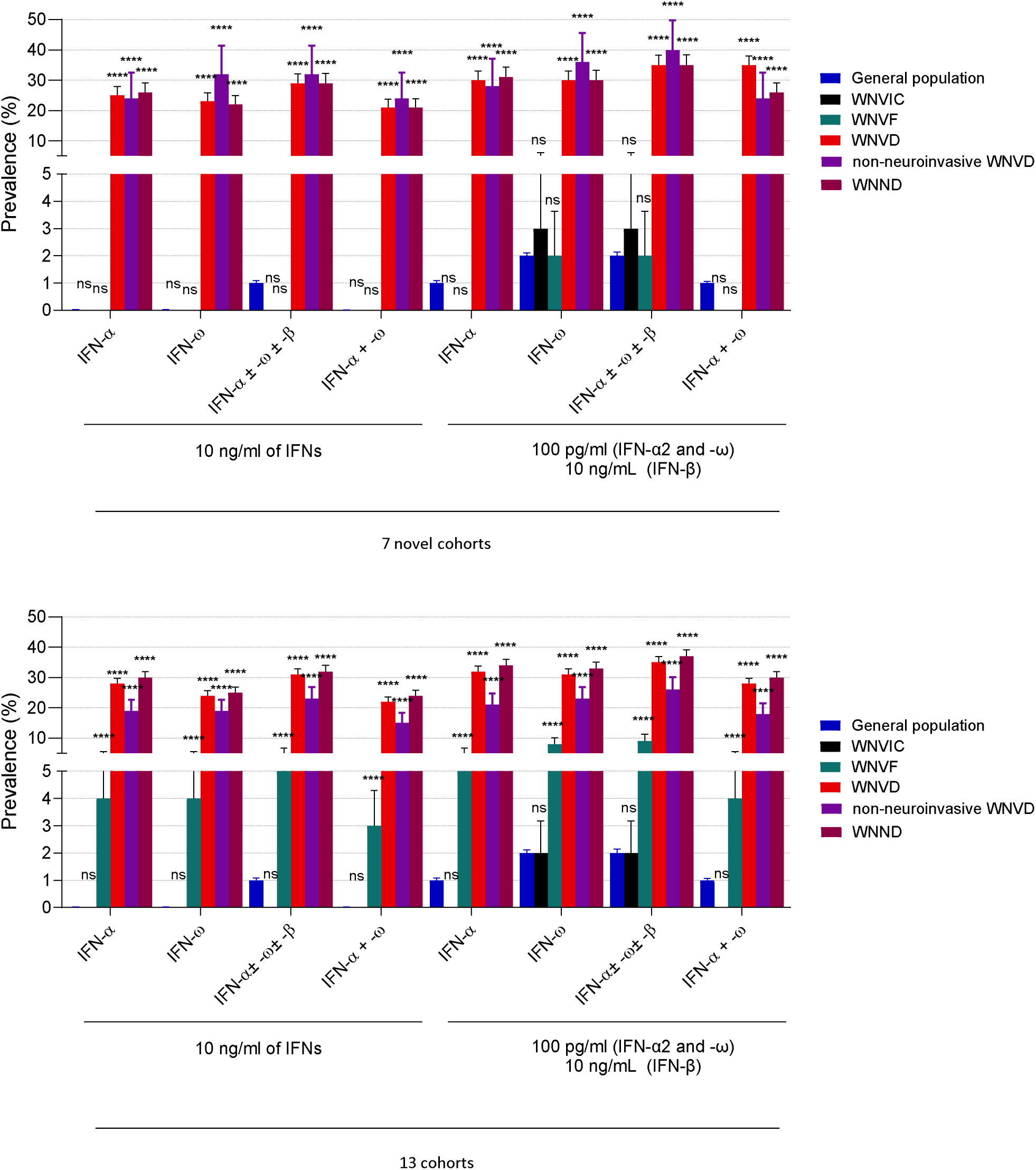
Frequency of auto-Abs against type I IFNs in the WNVIC, WNVF, and WNVD groups and the two WNVD subgroups (WNVD without evidence of neuroinvasive disease, and WNND) relative to the general population in the seven new cohorts and the overall study population consisting of all 13 cohorts. Horizontal bars represent the 95% CI limits. IFN-α: autoantibodies neutralizing IFN-α2 (regardless of effects on other IFNs); IFN-ω: autoantibodies neutralizing IFN-ω; IFN-α ± ω ± β: autoantibodies neutralizing IFN-α2 and/or IFN-ω and/or IFN-β; IFN-α + ω: autoantibodies neutralizing both IFN-α2 and IFN-ω. ns: non-significant; \*\*\*\**p* < 10^−4^.

**Figure 6.**
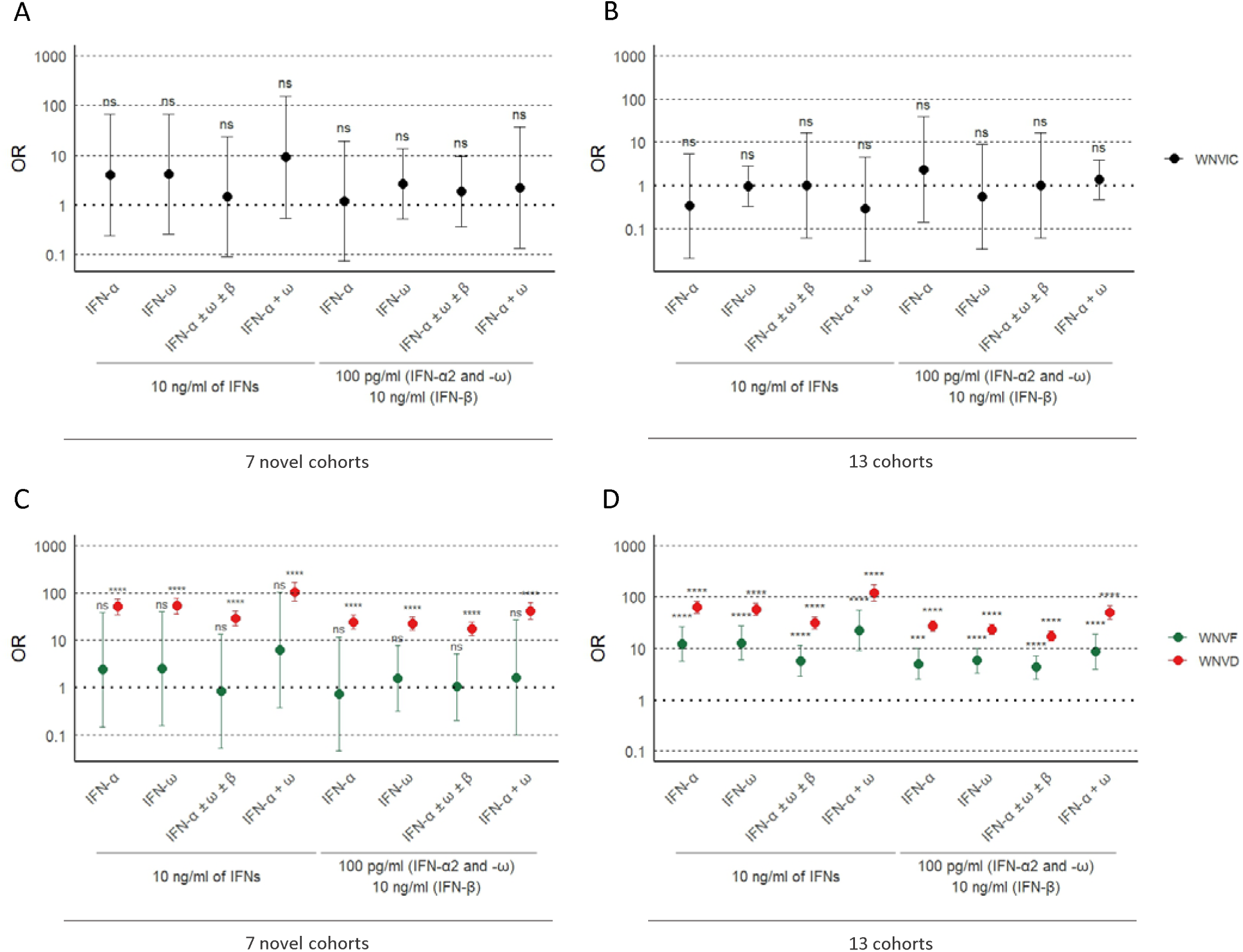
Odds ratios for the presence of auto-Abs in the WNV groups relative to the general population. Odds ratios (ORs) for the presence of auto-Abs in the WNVIC group relative to the general population, adjusted for age and sex with Firth’s bias-corrected logistic regression, in the seven new cohorts (A) and the overall study population consisting of 13 cohorts (B). ORs for the presence of auto-Abs in individuals with WNVF and WNVD in the seven new cohorts (C) and the overall study population consisting of 13 cohorts (D) relative to the general population, also adjusted for age and sex via logistic regression. Horizontal bars represent the 95% CI limits. IFN-α: autoantibodies neutralizing IFN-α2 (regardless of effects on other IFNs); IFN-ω: autoantibodies neutralizing IFN-ω; IFN-α ± ω ± β: autoantibodies neutralizing IFN-α2 and/or IFN-ω and/or IFN-β; IFN-α + ω: autoantibodies neutralizing both IFN-α2 and IFN-ω. Ns: non-significant; \*\*\*\**p* < 10^−4^.

**Table 3.** Risk of WNVF or WNVD in subjects carrying auto-Abs neutralizing specific sets of type I IFNs, relative to the general population, with adjustment for age and sex and risk of WNVD by age group.

| Anti-type I IFN auto-Ab<br>(amount of type I IFN<br>neutralized, in plasma<br>diluted 1:10) | WNV group | New cohort |  | Overall cohort |  |
| --- | --- | --- | --- | --- | --- |
|  |  | OR [95%CI] | p value | OR [95%CI] | p value |
| Anti-IFN-ω (100 pg/mL) | WNVF | 1.6 [0.3-7.8] | 0.6 | 5.8 [3.3-10.1] | $5.0 \times 10^{-7}$ |
| Anti-IFN-$\alpha 2$ (100<br>pg/mL) | WNVF | 0.7 [0.1-11.8] | 0.8 | 5 [2.3-9.9] | $1.5 \times 10^{-4}$ |
| Anti-IFN-$\alpha 2$ (100<br>pg/mL) and/or anti-IFN-ω (100 pg/mL)<br>and/or anti-IFN-β (10<br>ng/mL) | WNVF | 1.0 [0.2-5.2] | 1.0 | 4.3 [2.6-7.3] | $4.1 \times 10^{-6}$ |
| Anti-IFN-$\alpha 2$ (100<br>pg/mL) and anti-IFN-ω<br>(100 pg/mL) | WNVF | 1.6 [0.1-26.4] | 0.8 | 8.7 [4-19.1] | $2.1 \times 10^{-5}$ |
| Anti-IFN-ω (10 ng/mL) | WNVF | 2.5 [0.2-39.8] | 0.6 | 12.8 [6-27.5] | $1.2 \times 10^{-6}$ |
| Anti-IFN-$\alpha 2$ (10 ng/mL) | WNVF | 2.4 [0.1-39.1] | 0.6 | 12.1 [5.6-26.2] | $2 \times 10^{-6}$ |
| Anti-IFN-$\alpha 2$ (10 ng/mL)<br>and/or anti-IFN-ω (10<br>ng/mL) and/or anti-IFN-β (10 ng/mL) | WNVF | 0.8 [0.1-13.4] | 0.9 | 5.7 [2.9-11.3] | $5.3 \times 10^{-5}$ |
| Anti-IFN-$\alpha 2$ (10 ng/mL)<br>and anti-IFN-ω (10<br>ng/mL) | WNVF | 6.2 [0.4-101.9] | 0.3 | 22.5 [9-56.1] | $1.8 \times 10^{-6}$ |
| Anti-IFN-$\alpha 2$ (10 ng/mL)<br>and anti-IFN-ω (100<br>pg/mL) | WNVF | 3.0 [0.2-50.6] | 0.5 | 14.0 [5.7-34] | $6.1 \times 10^{-6}$ |
| Anti-IFN-ω (100 pg/mL) | WNVD (all) | 22.6 [16.2-31.2] | $< 10^{-16}$ | 23.4 [18.8-29.3] | $< 10^{-16}$ |
| | WNVD $\leq 65$ | 32 [17-59.9] | $< 10^{-16}$ | 26 [17-39.6] | $< 10^{-16}$ |
| | WNVD $> 65$ | 27.7 [18.3-41.9] | $< 10^{-16}$ | 26.9 [20.4-35.5] | $< 10^{-16}$ |
| Anti-IFN-$\alpha 2$ (100<br>pg/mL) | WNVD (all) | 24.5 [17.3-34.5] | $< 10^{-16}$ | 27.2 [21.5-34.4] | $< 10^{-16}$ |
| | WNVD $\leq 65$ | 89.2 [43.9-181.3] | $< 10^{-16}$ | 66.3 [38.1-115.4] | $< 10^{-16}$ |
| | WNVD $> 65$ | 21.8 [14.4-32.8] | $< 10^{-16}$ | 24.7 [18.9-32.4] | $< 10^{-16}$ |
| Anti-IFN-$\alpha 2$ (100<br>pg/mL) and/or anti-IFN-ω (100 pg/mL)<br>and/or anti-IFN-β (10<br>ng/mL) | WNVD (all) | 17.4 [12.7-23.7] | $< 10^{-16}$ | 17.4 [14.1-21.4] | $< 10^{-16}$ |
| | WNVD $\leq 65$ | 27.9 [16.3-47.8] | $< 10^{-16}$ | 19.2 [13.2-27.9] | $< 10^{-16}$ |
| | WNVD $> 65$ | 17.8 [11.9-26.4] | $< 10^{-16}$ | 19 [14.5-24.6] | $< 10^{-16}$ |
| Anti-IFN-$\alpha 2$ (100<br>pg/mL) and anti-IFN-ω<br>(100 pg/mL) | WNVD (all) | 41.7 [27.9-62.2] | $< 10^{-16}$ | 49.7 [37-67.1] | $< 10^{-16}$ |
| | WNVD $\leq 65$ | 426.4 [114.5-1587.9] | $< 10^{-16}$ | 300 [106.5-845] | $< 10^{-16}$ |
| | WNVD $> 65$ | 38.3 [24.3-60.2] | $< 10^{-16}$ | 40.5 [29.3-56] | $< 10^{-16}$ |
| Anti-IFN-ω (10 ng/mL) | WNVD (all) | 52.9 [36-77.5] | $< 10^{-16}$ | 57.1 [43.5-75] | $< 10^{-16}$ |
| | WNVD $\leq 65$ | 162 [75.2-348.8] | $< 10^{-16}$ | 138.9 [81.7-236] | $< 10^{-16}$ |
| | WNVD $> 65$ | 43.7 [28.3-67.4] | $< 10^{-16}$ | 42 [31-57] | $< 10^{-16}$ |
| Anti-IFN-$\alpha 2$ (10 ng/mL) | WNVD (all) | 51.1 [35-74.4] | $< 10^{-16}$ | 63.1 [48.6-81.8] | $< 10^{-16}$ |
| | WNVD $\leq 65$ | 300.2 [137.4-665.1] | $< 10^{-16}$ | 213.7 [117.6-388.6] | $< 10^{-16}$ |
| | WNVD $> 65$ | 37.4 [24.5-57.2] | $< 10^{-16}$ | 46.9 [35.3-62.3] | $< 10^{-16}$ |
| Anti-IFN-$\alpha 2$ (10 ng/mL)<br>and/or anti-IFN-ω (10<br>ng/mL) and/or anti-IFN-β (10 ng/mL) | WNVD (all) | 28.8 [20-41.4] | $< 10^{-16}$ | 31.2 [23.9-40.7] | $< 10^{-16}$ |
| | WNVD $\leq 65$ | 60.3 [30.1-120.8] | $< 10^{-16}$ | 44.4 [26.7-74] | $< 10^{-16}$ |
| | WNVD $> 65$ | 25.8 [16.8-39.9] | $< 10^{-165}$ | 29 [21.2-39.6] | $< 10^{-16}$ |
| Anti-IFN-$\alpha 2$ (10 ng/mL)<br>and anti-IFN-ω (10<br>ng/mL) | WNVD (all) | 104.6 [66.5-164.4] | $< 10^{-16}$ | 119.7 [83.8-171] | $< 10^{-16}$ |
| | WNVD $\leq 65$ | 548.2 [173.3-1733.9] | $< 10^{-16}$ | 538.2 [208.1-1391.8] | $< 10^{-16}$ |
| | WNVD $> 65$ | 80 [49.1-130.5] | $< 10^{-16}$ | 80.8 [55.4-117.8] | $< 10^{-16}$ |
| Anti-IFN-$\alpha 2$ (10 ng/mL)<br>and anti-IFN-ω (100<br>pg/mL) | WNVD (all) | 57.2 [34.1-95.8] | $< 10^{-16}$ | 77.5 [50.5-118.9] | $< 10^{-16}$ |
| | WNVD $\leq 65$ | 1730.9 [92.4-32443.2] | $2.2 \times 10^{-16}$ | 1517 [91.5-25181.1] | $< 10^{-16}$ |
| | WNVD $> 65$ | 45.2 [26.2-78.1] | $< 10^{-16}$ | 55.3 [35.7-85.5] | $< 10^{-16}$ |
| Anti-IFN-$\alpha 2$ (10 ng/mL)<br>and anti-IFN-ω (10<br>ng/mL) and anti-IFN-β<br>(10 ng/mL) | WNVD (all) | 347.7 [18.9-6381.4] | $5.9 \times 10^{-9}$ | 173.0 [10.0-3002.8] | $4.9 \times 10^{-10}$ |
| | WNVD $\leq 65$ | 200.2 [9.4-4270.8] | $7.9 \times 10^{-5}$ | 74.0 [3.5-1559.6] | $9 \times 10^{-4}$ |
| | WNVD $> 65$ | 177.2 [9.0-3497.7] | $1.1 \times 10^{-5}$ | 116.8 [6.6-2063.7] | $1.1 \times 10^{-7}$ |

WNVF: West Nile virus fever; WNVD: West Nile virus disease.
$\leq 65$ and $> 65$ indicate age cut-offs, in years; anti-IFN-ω and anti-IFN- $\alpha 2$ indicate auto-Abs neutralizing IFN-ω or IFN- $\alpha 2$ , respectively, regardless of their effects on other type I IFNs

### Risk of WNVF conferred by the auto-Abs in the new and total cohorts

The criteria for assignment to the WNV disease were identical for this and the initial study but, for several of the newly recruited cases, we obtained access to longitudinal documentation, making it possible to distinguish accurately between true WNVF cases (that is, individuals never hospitalized during the course of WNV infection) and patients initially assigned to the WNVF group but subsequently displaying progression to more severe disease. A longitudinal analysis of the clinical data showed that 17/78 [22%] of the patients initially referred to us for WNVF were later hospitalized (and were therefore reassigned to the WNVD group), with clinical signs of neuroinvasive disease reported in 12/17 (71%) cases. Strikingly, samples from one of these 17 patients (6%) neutralized both high and low concentrations of IFN-α2 and samples from another patient (6%) neutralized both high and low concentrations of IFN-ω. This two-step course of the disease is reminiscent of that observed in severe SARS-CoV-2 infection, for which a pathogenetic model has been proposed (37). These observations also highlight the need for consistent definitions of WNV disease categories, particularly for WNVF, as considerable heterogeneity is observed between studies (38,39,40), with the definition of WNVF ranging from self-reported, mild clinical disease (14) to severe clinical disease requiring hospitalization in individuals not undergoing lumbar puncture (41). This variability of the definition of WNVF is reflected in the definitions used at different clinical centers. Moreover, individuals with mild clinical disease rarely undergo diagnostic testing for WNV, even in regions with endemic WNV circulation, suggesting that most of the samples collected from individuals with WNVF reflect the most severe clinical cases, for which medical attention is sought. Overall, the study of seven new cohorts of patients with WNV infection modulates our previous findings, suggesting that the risk of mild WNVF is increased little, if at all, by the presence of auto-Abs neutralizing type I IFNs, at least under the experimental conditions used here to test for type I IFN-neutralizing activity in patient samples.

### Risk of WNVD in individuals with auto-Abs neutralizing type I IFNs

The prevalence of auto-Abs, both overall and in various combinations, was significantly higher in WNVD patients than in the general population. The presence of auto-Abs neutralizing at least low concentrations of IFN-α2 and/or IFN-ω was associated with a higher risk of WNVD in both the new cohorts (OR = 18.7; 95% CI: 13.8-25.4, *p* < 10^−16^) (Fig. 5 and 6C, table 3), and in the overall study population consisting of the new and previously studied cohorts analyzed together (OR = 18.9; 95% CI: 15.4-23.1, *p* < 10^−16^) (Fig. 5 and 6D, table 3). A combination of auto-Abs neutralizing at least low concentrations of both IFN-α2 and IFN-ω further increased the risk of WNVD in the new cohorts (OR = 41.7; 95% CI: 28.0-62.3, *p* < 10^−16^) (Fig. 5 and 6C, table 3) and in the overall study population consisting of the new and previously studied cohorts analyzed together (OR = 49.7; 95% CI: 36.9-67.1, *p* < 10^−16^) (Fig. 5 and 6D, table 3). The risk of clinical disease was higher in individuals carrying auto-Abs able to neutralize high concentrations (10 ng/mL) of IFN-α2, IFN-β, or IFN-ω. The presence of auto-Abs neutralizing high concentrations of IFN-α2 only, IFN-ω only, IFN-β only or combinations of IFN-α2 and/or IFN-β and/or IFN-ω resulted in a ∼4 to ∼80 times higher risk of severe WNVD (table 3). The risk was higher in individuals with auto-Abs neutralizing a combination of high concentrations (10 ng/mL) of both IFN-α2 and IFN-ω, with a ∼65 to 120 times increase in the risk (OR = 105; 95% CI: 67-164, *p* < 10^−16^, and OR = 120; 95% CI: 84-171, *p* < 10^−16^) for auto-Abs neutralizing high concentrations of both IFN-α2 and IFN-ω regardless of the presence of auto-Abs neutralizing IFN-β, in the new and total cohorts, respectively (Fig. 6C-D, table 3). Individuals with a combination of auto-Abs neutralizing high concentrations (10 ng/mL) of IFN-α2, IFN-β and IFN-ω had the highest risk of severe clinical disease, with a >150-fold increase in the risk of WNVD (OR = 348; 95% CI: 19-6381, *p* = 5.9 × 10^−9^; and OR = 173; 95% CI: 10-3003, *p* = 4.9 × 10^−10^ in the new and total cohorts, respectively) (table 3).

### Risk of neuroinvasive disease in individuals with auto-Abs neutralizing at least low concentrations of type I IFNs

Among WNVD patients, the prevalence of auto-Abs, overall and in various combinations, was significantly higher in the subgroup of individuals with neuroinvasive disease than in the general population. The presence of auto-Abs neutralizing at least low concentrations of IFN-α2 and/or IFN-ω was associated with a higher risk of neuroinvasive disease, encephalitis in particular, in both the new cohorts (OR = 18.1; 95% CI: 13.1-25.1, *p* < 10^−16^ and OR = 18.8; 95% CI: 13.1-27, *p* < 10^−16^ for neuroinvasive disease and encephalitis, respectively), and in the overall study population consisting of the new and previously studied cohorts analyzed together (OR = 20.2; 95% CI: 16.2-25, *p* < 10^−16^ and OR = 17.7; 95% CI: 13.8-22.8, *p* < 10^−16^ for neuroinvasive disease and encephalitis, respectively) (Figs. 5 and 7A-D, table S3). A combination of auto-Abs neutralizing at least low concentrations of both IFN-α2 and IFN-ω further increased the risk of neuroinvasive disease, encephalitis in particular, in both the new cohorts (OR = 42.2; 95% CI: 27.8-63.9 and OR = 46.4; 95% CI: 29.8-72.2, *p* < 10^−16^, *p* < 10^−16^ for neuroinvasive disease and encephalitis, respectively) and in the overall study population (OR = 53.6; 95% CI: 39.4-72.9, *p* < 10^−16^ and OR = 46.4; 95% CI: 33.1-65, *p* < 10^−16^ for neuroinvasive disease and encephalitis, respectively) (Fig. 7A-D, table S3).

**Figure 7.**
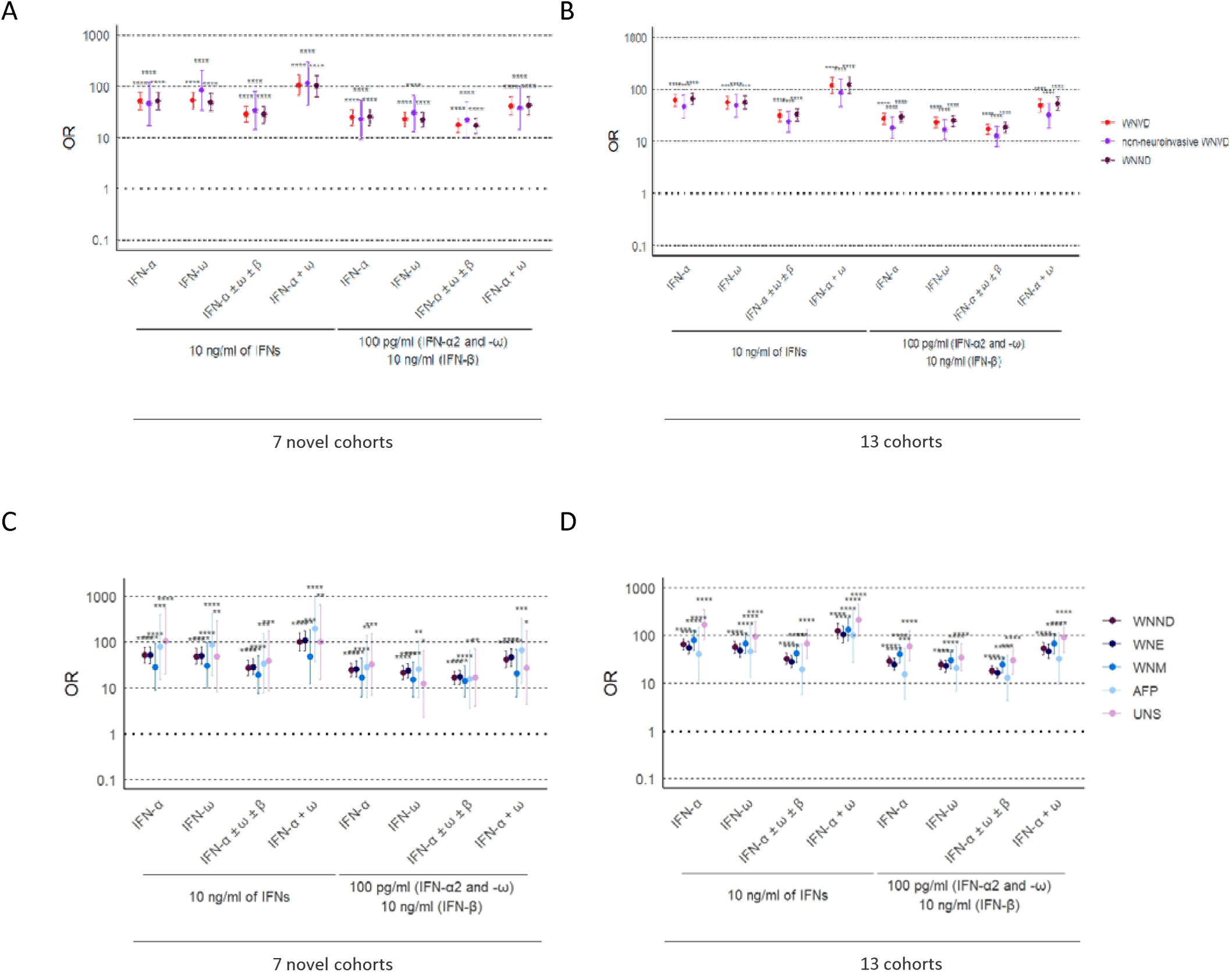
Odds ratios for the presence of auto-Abs in the WNVD group relative to the general population. Odds ratios (ORs) for the presence of auto-Abs in the WNVD group and the two WNVD subgroups (WNVD without evidence of neuroinvasive disease, and WNV neuroinvasive disease; WNND) relative to the general population, adjusted for age and sex with Firth’s bias-corrected logistic regression, in the seven new cohorts (A) and the overall study population consisting of 13 cohorts (B). Odds ratios (ORs) for the presence of auto-Abs in the WNND group and the four WNND subgroups (encephalitis; WNE, meningitis; WNM, acute flaccid paralysis; AFP, and unspecified neurological syndrome; UNS) in the seven new cohorts (C) and in overall study population consisting of 13 cohorts (D) relative to the general population, also adjusted for age and sex via logistic regression. Horizontal bars represent the 95% CI limits. IFN-α: autoantibodies neutralizing IFN-α2 (regardless of effects on other IFNs); IFN-ω: autoantibodies neutralizing IFN-ω; IFN-α ± ω ± β: autoantibodies neutralizing IFN-α2 and/or IFN-ω and/or IFN-β; IFN-α + ω: autoantibodies neutralizing both IFN-α2 and IFN-ω. ns: non-significant; \*\*\*\**p* < 10^−4^.

### Risk of neuroinvasive disease in individuals with auto-Abs neutralizing high concentrations of type I IFNs

The presence of auto-Abs neutralizing high concentrations of IFN-α2 only, IFN-ω only, IFN-β only or combinations of IFN-α2 and/or IFN-β and/or IFN-ω resulted in a ∼3 to ∼60 times higher risk of severe WNVD neuroinvasive disease, encephalitis in particular (Fig. 7A-D, table S3). The presence of auto-Abs neutralizing a combination of high concentrations (10 ng/mL) of both IFN-α2 and IFN-ω resulted in a ∼60 to >200 times increase in the risk of neuroinvasive disease, encephalitis in particular, in both the new cohorts (OR = 101.3; 95% CI: 63.3-162.1, *p* < 10^−16^ and OR = 106.7; 95% CI: 65-175, *p* < 10^−16^ for Abs neutralizing high concentrations of both IFN-α2 and IFN-ω regardless of the presence of auto-Abs neutralizing IFN-β, for neuroinvasive disease and encephalitis, respectively), and the overall study population (OR = 123.5; 95% CI: 85.6-178.3, *p* < 10^−16^ and OR = 105.8; 95% CI: 71.1-157.3, *p* < 10^−16^ for Abs neutralizing high concentrations of both IFN-α2 and IFN-ω regardless of the presence of auto-Abs neutralizing IFN-β, for neuroinvasive disease and encephalitis, respectively) (Fig. 7A-D, table S3). Individuals with a combination of auto-Abs neutralizing high concentrations (10 ng/mL) of IFN-α2 and IFN-β and IFN-ω had the highest risk of neuroinvasive disease, encephalitis in particular, in the new cohorts (OR = 243.9; 95% CI: 12.5-4744.1, *p* = 2.87 × 10^−6^ and OR = 218.5; 95% CI: 10.3-4629.5, 6.04 × 10^−5^ for neuroinvasive disease and encephalitis, respectively) and the overall cohort (OR = 141.9; 95% CI: 7.9-2535.1, *p* = 1.08 × 10^−7^ and OR = 127.9; 95% CI: 6.8-2413.9, *p* = 5.11 × 10^−6^ for neuroinvasive disease and encephalitis, respectively) (table S3).

### Risk of WNVD due to auto-Abs neutralizing type I IFNs in subjects **≤** 65 years old

The calculated risk of WNVD conferred by the auto-Abs was greatest in subjects ≤ 65 years old, for all combinations of auto-Abs neutralizing low, intermediate or high concentrations of the three type I IFNs tested (Fig. 8A-D, table 3). In subjects ≤ 65 years old, the presence of auto-Abs neutralizing high concentrations (10 ng/mL) of IFN-α2 combined with auto-Abs neutralizing high (10 ng/mL) or low (100 pg/mL) concentrations of IFN-ω resulted in a ∼500 to ∼1500 increase in the risk of WNVD in the new cohorts (OR = 548.2; 95% CI: 173.4-1733.9, *p* < 10^−16^ for a combination of auto-Abs neutralizing high concentrations of both IFN-α2 and IFN-ω; and OR = 1730.9; 95% CI: 92.4-32443.2, *p* = 2.22 × 10^−16^ for auto-Abs neutralizing high concentrations of IFN-α2 and any concentration tested [high or low] of IFN-ω) (Fig. 8A-D, table 3) and the overall study population (OR = 538.2; 95% CI: 208.1-1391.8, *p* < 10^−16^ for auto-Abs neutralizing high concentrations of both IFN-α2 and IFN-ω; and OR = 1517.7; 95% CI: 91.5-25181.1, *p* < 10^−16^ for auto-Abs neutralizing high concentrations of IFN-α2 and any concentration tested [high or low] of IFN-ω) (Fig. 8A-D, table 3). The calculated risk was similarly high in the subgroup of patients with neuroinvasive disease in the new cohorts (OR = 442; 95% CI: 128.6-1518.6, p = 2.45 × 10^−14^ for a combination of auto-Abs neutralizing high concentrations of both IFN-α2 and IFN-ω; and OR = 1616.4; 95% CI: 84.1-31074.1, *p* = 2.31 × 10^−13^ for auto-Abs neutralizing high concentrations of IFN-α2 and any concentration tested [high or low] of IFN-ω) and the total cohort (OR = 602; 95% CI: 224.2-1616.2, *p* < 10^−16^ for a combination of auto-Abs neutralizing high concentrations of both IFN-α2 and IFN-ω; and OR = 1841.1; 95% CI: 109.5-30940.6, *p* < 10^−16^ for auto-Abs neutralizing high concentrations of IFN-α2 and any concentration tested [high or low] of IFN-ω) (table S3). In the subgroup of patients with encephalitis, the most severe manifestation of WNV infection, auto-Abs neutralizing at least 100 pg/mL of both IFN-α2 and IFN-ω conferred a ∼400 to ∼700 times increase in risk (OR = 705.6; 95% CI: 138.6-3592.6, *p* = 3.3 × 10^−15^ and OR = 420.7; 95% CI: 128.5-1377.3, *p* < 10^−16^ for the new and total cohorts, respectively), whereas auto-Abs neutralizing both high concentrations of IFN-α2 and any concentration tested (high or low) of IFN-ω were associated with the highest risk, > 2000 times higher than for the reference group (OR = 2597.5; 95% CI: 117.1-57597.3, *p* = 3.93 × 10^−12^ and OR = 2218.4; 95% CI: 125.1-39337.7, *p* < 10^−16^ for the novel and total cohorts, respectively) (table S3).

**Figure 8.**
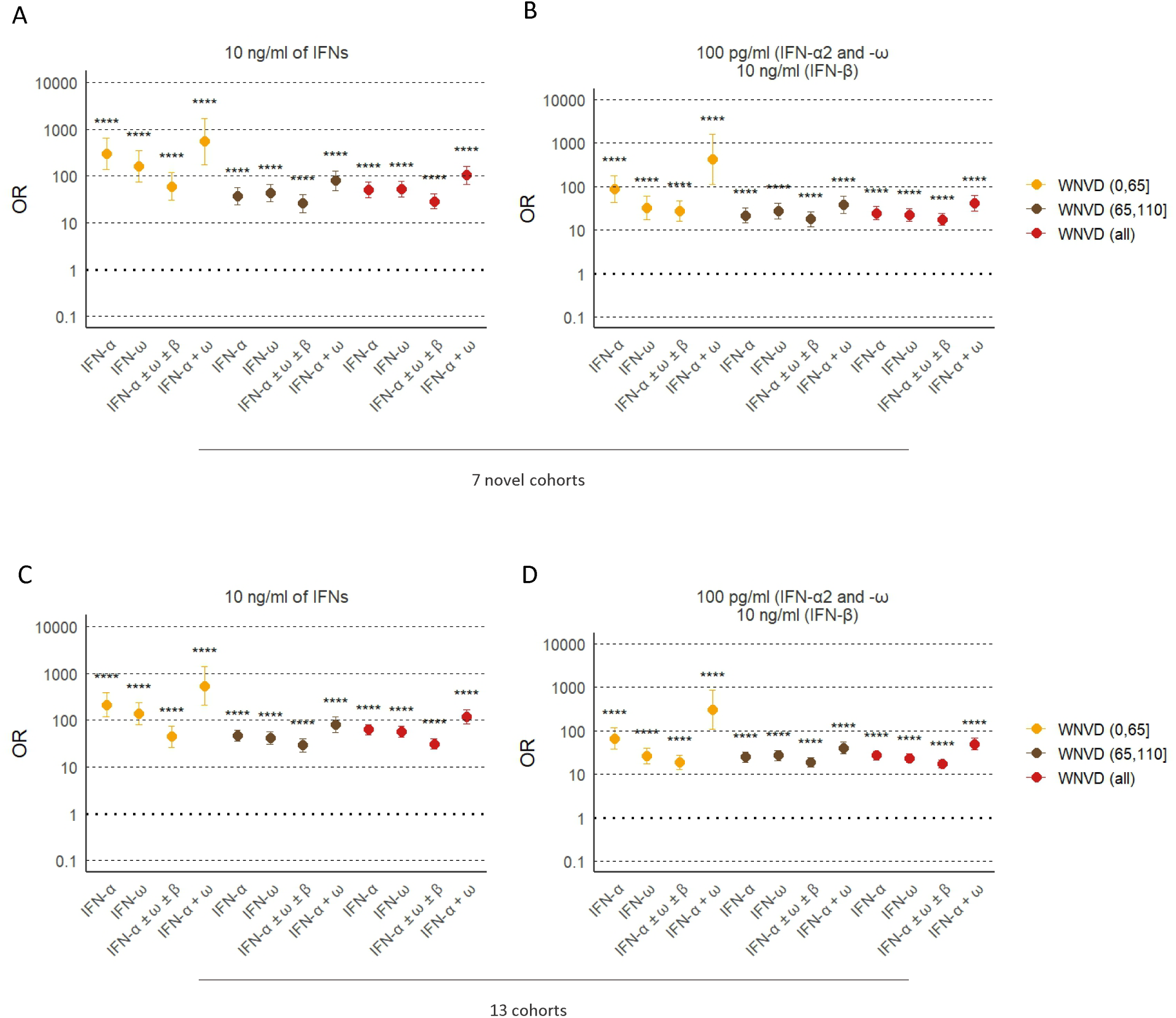
Odds ratios for the presence of auto-Abs in the WNV groups relative to the general population, by age group. ORs for the presence of auto-Abs in patients with WNVD relative to the general population, as determined by logistic regression, stratified for age group, for auto-Abs neutralizing different combinations of high concentrations (A) and low concentrations (B) of type I IFNs in the seven new cohorts, and for auto-Abs neutralizing different combinations of high concentrations (C) and low concentrations (D) of type I IFNs in the overall study population consisting of 13 cohorts. ORs were calculated separately for patients aged ≤65 and >65 years. Horizontal bars represent the 95% CI limits. IFN-α: autoantibodies neutralizing IFN-α2 (regardless of effects on other IFNs); IFN-ω: autoantibodies neutralizing IFN-ω; IFN-α ± ω ± β: autoantibodies neutralizing IFN-α2 and/or IFN-ω and/or IFN-β; IFN-α + ω: autoantibodies neutralizing both IFN-α2 and IFN-ω. ns: non-significant; \*\*\*\**p* < 10^−4^.

### Risk of WNVD due to auto-Abs neutralizing type I IFNs in subjects > 65 years old

In subjects > 65 years old, a combination of auto-Abs neutralizing a combination of high concentrations of both IFN-α2 and IFN-ω, regardless of the neutralization of IFN-β, resulted in a ∼80 times higher risk of WNVD (OR = 80.0; 95% CI: 49.1-130.5, *p* < 10^−16^ and OR = 80.8; 95% CI: 55.4-117.8, *p* < 10^−16^, in the new and total cohorts, respectively) (Fig. 8A-D), neuroinvasive disease (OR = 82.9; 95% CI: 50.2-137, *p* < 10^−16^ and OR = 84.6; 95% CI: 57.6-124.3, *p* < 10^−16^ in the new and total cohorts, respectively), and encephalitis in particular (OR = 86.9; 95% CI: 51.5-146.8, *p* < 10^−16^ and OR = 76.1; 95% CI: 50.3-115.1, *p* < 10^−16^ in the new and total cohorts, respectively) (table S3). A combination of auto-Abs neutralizing high concentrations of the three auto-Abs tested conferred the highest risk in this age group, with a >100 times increase in the risk of WNVD and neuroinvasive disease (table 3). The ORs calculated by age group for all combinations of auto-Abs are reported in tables 3 and S3. Overall, the presence of auto-Abs neutralizing type I IFNs confers a very high risk of WNVD in all age groups. The biological and medical impact of various combinations of auto-Abs was considerable in both the under- and over-65s. The lower ORs calculated for individuals aged 65 years or older reflect both a higher baseline risk of severe disease in the absence of auto-Abs, and the higher baseline prevalence of auto-Abs in the elderly (42). These results are consistent with previous reports on anti-type I IFN auto-Abs in individuals < 65 or 70 years old with hypoxemic COVID-19 pneumonia (42), critical influenza pneumonia (43), or WNVD (20). Overall, the data for the seven new cohorts of WNVD patients from five countries confirm the strong effect of auto-Abs neutralizing type I IFNs on the risk of developing severe disease following exposure to WNV, and demonstrate that impaired type I IFN immunity is a major determinant of WNVD and neuroinvasive disease. Auto-Abs underlie life-threatening WNVD in a significant proportion of cases, considerably increasing the likelihood of neuroinvasive disease and encephalitis in particular.

## Discussion

These findings extend those of our previous report (20) and confirm that auto-Abs neutralizing type I IFNs underlie almost half of all cases of WNV encephalitis. These auto-Abs have now been found in 13 unrelated cohorts from nine centers in five countries in Europe, the Middle East, and North America, in samples collected from 2002 to 2024. When analyzed collectively, the ORs for encephalitis were found to be very high, ranging from 20 for auto-Abs neutralizing low concentrations of IFN-α2 and/or IFN-ω to >2000 for auto-Abs neutralizing high concentrations of IFN-α2 together with IFN-ω in the under-65s. The proportion of cases with neuroinvasive disease carrying auto-Abs neutralizing one or more type I IFN was also very high, and equally so across cohorts, with a mean frequency of 38% and a range of 17% to 50% of cases. These variations may depend on differences in disease group assignment, suggesting that consistent definitions of disease severity and neurological involvement and follow-up data for infected individuals are required.

Such high ORs and proportions are unprecedented for human infectious diseases (44,45). The ORs were calculated by comparing patients with WNV encephalitis relative to individuals untested for WNV infection in a large sample of the general population. In our cohort, the proportion of individuals with silent WNV infection carrying the auto-Abs did not significantly differ from that in the general population. Yet, our sample of WNVIC is much smaller (146 WNVIC vs >10,000 individuals of the corresponding age groups) and includes only 14 subjects > 65 years old, none of whom carried the auto-Abs. It is reasonable to predict that the proportion of individuals carrying the auto-Abs among those with silent WNV infection, selected as appropriate controls on the basis of their resistance to severe disease, could be even smaller than that in the general population, in particular among individuals > 65 years old, resulting in higher ORs for WNVD and encephalitis. This is also suggested by our observation that, in the new cohorts, no samples from WNVIC and mild WNVF patients neutralized IFN-α2, and there were no subjects with auto-Abs neutralizing more than one type I IFN, while 1.4% individuals in the general population carry auto-Abs neutralizing IFN-α2 and 0.6% carry auto-Abs neutralizing two or more type I IFNs (23).

The cells infected with WNV in which type I IFN activity is blocked by these auto-Abs, resulting in encephalitis, remain to be identified (22). Candidates include leukocytes, cells at the blood-brain barrier, and cells within the central nervous system (22). This study has important clinical implications. People at risk of having auto-Abs against type I IFNs, including individuals with a history of autoimmunity or viral disease, or over the age of 70 years, should be screened. Patients with auto-Abs neutralizing type I IFNs should be considered at risk and should avoid inhabiting or traveling to areas in which WNV is endemic. If this cannot be avoided or if they are outdoor workers living in endemic areas (46,47), they should protect themselves against mosquitoes and, if bitten, should seek medical attention before the possible development of clinical disease. Patients hospitalized for WNV encephalitis should be tested for auto-Abs, as should patients diagnosed with WNV infection in other contexts. Treatment with a type I IFN not neutralized by the auto-Abs, such as IFN-beta (48) or, in the future, with decoys that neutralize the auto-Abs but not the IFNs and their receptors (32), or with CAR-T cells (49), may also be considered.

In 2020, we concluded that auto-Abs against type I IFNs were causal for critical COVID-19, for several individually indirect but collectively compelling reasons (31), as summarized by Manry et al. (42). This interpretation was supported by the longitudinal follow-up of a Swiss cohort of patients living with human immunodeficiency virus (HIV) infection, which showed that these auto-Abs occur before severe viral diseases and that they later diversify and persist for life (50). The production of auto-Abs against type I IFNs results from a failure of tolerance to self, which can be driven by monogenic IEI affecting central tolerance in the thymus (51,52,53,54,55,56,57,58,59,60,61,62). Once these auto-Abs appear, they do not disappear.

They can underlie at least three severe viral respiratory diseases — COVID-19, influenza, and MERS — in 5-20% of cases (23,31,43,63,64). We show here that they are the major determinants of WNV encephalitis in North America, Europe, and the Middle East. They also underlie 10% of cases of TBE (26), and the rarer cases of encephalitis due to the Powassan and Usutu viruses (27). They were also found in the most severe case of Ross River disease, which is caused by an alphavirus (27). They should be sought in patients with other types of encephalitis, particularly due to arboviruses, which are typically harmless in most infected individuals but may cause severe disease in a minority of cases. There are over 150 known human-pathogenic arboviruses and studies to assess the contribution of auto-Abs against type I IFNs to the pathogenesis of each arboviral disease are warranted.

## Materials and Methods

### Patients

We enrolled an international cohort of 318 individuals aged 10 to 99 years with documented WNV infection, 65.1% of whom were male and 33.9% female, living in Austria, Hungary, Israel, Italy, or the USA (Fig. 1 and S1), and a cohort of 663 individuals described in a previous study (20), living in Hungary, Italy, or the USA. In the new cohorts, neither sex nor age was reported for 2/318 individuals, age was not reported for 2/318 individuals, and sex was not reported for 1/318 individuals. therein total, there were 981 individuals aged 9 to 99 years with documented WNV infection, 64.7% of whom were male and 34.9% female, living in Austria, Hungary, Israel, Italy, or the USA (Fig. S5). For all the enrolled individuals from the new and previously described cohorts, written informed consent was obtained in the country of residence of each patient, unless samples and anonymized medical information could be processed without prior written informed consent, in accordance with local regulations and with institutional review board (IRB) approval. The Center for Virology at the Medical University of Vienna, Austria, serves as the national reference center for arboviruses, and has the legal permission to process anonymized patient information on notifiable viral diseases. WNV infection was diagnosed on the basis of the serological demonstration of WNV-specific IgM or seroconversion to IgG, WNV neutralization assays (29), and/or RT-PCR on serum, plasma or cerebrospinal fluid samples. Individuals were stratified according to the presence and/or severity of clinical manifestations, as defined by the need for hospitalization. Life-threatening WNV disease (WNVD) was defined as WNV infection requiring hospitalization. WNV fever (WNVF) was defined as WNV infection not requiring hospitalization in patients reporting a febrile illness requiring outpatient care. The WNV-infected controls (WNVIC) were blood donors with documented WNV infection, diagnosed on the basis of the detection of WNV RNA in blood during screening at the time of blood donation, who remained asymptomatic or paucisymptomatic (headache) during follow-up. WNVD patients included individuals with confirmed neurological disease (WNV neuroinvasive disease) and individuals without clinical evidence of neuroinvasive disease. The individuals in the neuroinvasive disease group were reported to have encephalitis (fever, acute signs of central or peripheral neurologic dysfunction, including altered mental status and neurological deficits), meningitis (fever, pleocytosis, headache, nuchal rigidity), acute flaccid paralysis (poliomyelitis-like syndrome or Guillain-Barré-like syndrome) or other neurological syndromes. The experiments were conducted in Italy, France and the USA, in accordance with local regulations and guidance from the Italian national data protection authority, the French Ethics Committee (Comité de Protection des Personnes), the French National Agency for Medicine and Health Product Safety, the *Institut National de la Sant*é et de la Recherche Médicale in Paris, France, and with the approval of the IRB of the Italian institutions (San Matteo Research Hospital in Pavia [*Comitato Etico Territoriale Lombardia 6 - Policlinico San Matteo*], the University Hospital of Padua, the University Hospital of Bologna, Amedeo di Savoia Hospital-ASL Città in Torino in Turin), the Medical University of Vienna in Austria, the Tel-Aviv Sourasky Medical Center in Israel, the National Public Health Center in Budapest, and the Rockefeller University in New York, USA, respectively.

### ELISA

Enzyme-linked immunosorbent assays (ELISA) were performed as previously described (30). In brief, 96-well ELISA plates (MaxiSorp; Thermo Fisher Scientific) were coated by overnight incubation at 4°C with 1 μg/mL rhIFN-α (ref. number 130-108-984; Miltenyi Biotec), rhIFN-ω (ref. number 300-02J; Peprotech), or rhIFN-β (ref. number 300-02BC; Peprotech). The plates were washed (PBS/ 0.005% Tween), blocked by incubation with the same buffer supplemented with 2% BSA, washed, and incubated with 1:50 dilutions of plasma samples from the patients or positive and negative controls for 2 h at room temperature. Each sample was tested once. Plates were thoroughly washed (PBS/0.005% Tween) and horseradish peroxidase (HRP)– conjugated Fc-specific IgG fractions from polyclonal goat antiserum against human IgG (Nordic Immunological Laboratories) were added to a final concentration of 1 μg/mL. Plates were incubated for 1 h at room temperature and washed. The substrate was added, and optical density (OD) was measured (450 nm). All the incubation steps were performed with gentle shaking (600 rpm).

### Luciferase reporter assay

The blocking activity of anti-IFN-α2, anti-IFN-ω, and anti-IFN-β auto-Abs was determined with a luciferase reporter assay, as previously described (23). Briefly, HEK293T cells were transfected with a plasmid encoding the firefly luciferase gene under the control of the human ISRE promoter in the pGL4.45 backbone and a plasmid constitutively expressing the *Renilla* luciferase as a control for transfection (pRL-SV40). Cells were transfected in the presence of the X-tremeGene9 transfection reagent (ref. number 6365779001; Sigma-Aldrich). After 24 h, cells in Dulbecco’s modified Eagle medium (DMEM; Thermo Fisher Scientific) supplemented with 2% fetal calf serum (FCS) and 10% control or patient serum/plasma/whole blood (after heat inactivation at 56°C, for 20 min) were either left unstimulated or were stimulated with unglycosylated rhIFN-α2 (ref. number 130-108-984, Miltenyi Biotec), unglycosylated rhIFN-ω (ref. number 300-02J, Peprotech) at a concentration of 10 ng/mL or 100 pg/mL, or glycosylated rhIFN-β (ref. number 300-02BC, Peprotech) at a concentration of 10 or 1 ng/mL for 16 h at 37°C under an atmosphere containing 5% CO_2_. Finally, the cells were lysed by incubation with a lysis buffer (provided in ref. number E1980, Promega) for 20 min at room temperature and luciferase levels were measured with the Dual-Luciferase Reporter 1000 assay system (ref. number E1980, Promega) according to the manufacturer’s protocol. Luminescence intensity was measured with a VICTOR-X Multilabel Plate Reader (PerkinElmer Life Sciences). Firefly luciferase activity values were normalized against *Renilla* luciferase activity values. The resulting values (luciferase induction) were then normalized against the median level of induction for non-neutralizing samples and expressed as a percentage (relative luciferase activity (RLA) ratio, %). Samples were considered to have neutralizing activity if the RLA ratio was below 15% of the median value for controls tested on the same day.

### Statistical analysis

Odds ratios and *p* values for the effect of auto-Abs neutralizing each type I IFN in WNV patients relative to healthy individuals from the general population, adjusted for age in years and sex, were estimated by means of Firth’s bias corrected logistic regression, as implemented in the logistf package of R software. Where relevant, statistical test results are indicated in the corresponding figures. ns: not significant, \**p<*0.05, \*\**p<*0.01, \*\*\**p<*0.001, \*\*\*\**p<*0.0001.

## Abbreviations

Auto-Abs: autoantibodies
CI: confidence interval
COVID-19: coronavirus disease 2019
ELISA: enzyme-linked immunosorbent assay
HIV: human immunodeficiency virus
IEI: inborn error(s) of immunity
IFN: interferon
Ig: immunoglobulin
MERS: Middle East respiratory syndrome
NAAT: nucleic acid amplification test
OD: optical density
OR: odds ratio
POWV: Powassan virus
RRV: Ross River virus
RT-PCR: reverse transcription polymerase chain reaction
SARS-CoV-2: severe acute respiratory syndrome coronavirus 2
SD: standard deviation
TBE: tick-borne encephalitis
USUV: Usutu virus
WNV: West Nile virus
WNVD: West Nile virus disease
WNVF: West Nile virus fever
WNVIC: West Nile virus-infected controls
YFV-17D: yellow fever virus vaccine strain 17D

## Data availability statement

All data supporting the findings of this study are available within the main text and supplemental material and from the corresponding authors upon request.

## Acknowledgments

We thank the patients and their families for participating in our research. We thank all members of both branches of the Laboratory of Human Genetics of Infectious Diseases for discussions and technical and administrative support. A.Borghesi wishes to thank Giampaolo Merlini, University of Pavia, for thoughtful discussions on the methods for anti-cytokine auto-Ab discovery in samples from patients with bacterial and viral infections. We thank Lazaro Lorenzo, Laboratory of Human Genetics of Infectious Diseases, Necker Branch, Institut National de la Santé et de la Recherche Médicale (INSERM) U1163, Necker Hospital for Sick Children, Paris, France, EU.

## Funding

The Laboratory of Human Genetics of Infectious Diseases is supported by the Howard Hughes Medical Institute, The Rockefeller University, the St. Giles Foundation, the Stavros Niarchos Foundation (SNF) as part of its grant to the SNF Institute for Global Infectious Disease Research at The Rockefeller University, the National Institutes of Health (NIH) (R01AI163029), the National Center for Advancing Translational Sciences (NCATS), the NIH Clinical and Translational Science Award (CTSA) program (UL1TR001866), the French *Agence Nationale de la Recherche* (ANR) under the France 2030 program (ANR-10-IAHU-01), the HORIZON-HLTH-2024-DISEASE-08-20 program under GA 101191725 (InFlaMe), the ANRS projects DéméléJEV (ANRS0629) and LSDengue (ANRS-23-PEPR-MIE-0007), the Integrative Biology of Emerging Infectious Diseases Laboratory of Excellence (ANR-10-LABX-62-IBEID), the French Foundation for Medical Research (FRM) (EQU202503020018), ANR GENVIR (ANR-20-CE93-003), and ANR AI2D (ANR-22-CE15-0046) projects, the HORIZON-HLTH-2021-DISEASE-04 program under grant agreement 101057100 (UNDINE), the ANR-RHU COVIFERON Program (ANR-21-RHUS-0008), the Square Foundation, *Grandir - Fonds de solidarité pour l’enfance*, the *Fondation du Souffle*, the SCOR Corporate Foundation for Science, the Battersea & Bowery Advisory Group, William E. Ford, General Atlantic’s Chairman and Chief Executive Officer, Gabriel Caillaux, General Atlantic’s Co-President, Managing Director and Head of Business in EMEA, and the General Atlantic Foundation, the French Ministry of Higher Education, Research, and Innovation (MESRI-COVID-19), *Institut National de la Santé et de la Recherche Médicale* (INSERM), REACTing-INSERM, Paris Cité University, and the Imagine Institute. P.B. was supported by a “*Poste CCA-INSERM-Bettencourt*” (with support from the Bettencourt-Schueller Foundation), and the FRM (EA20170638020). A.G. was supported by a French National Agency for Research grant as part of the “*Investissement d’Avenir*” program (ANR-10-LABX-62-01). The Host-pathogen group (A. Borghesi), *Fondazione IRCCS Policlinico San Matteo*, Pavia, Italy, is supported by grants from the Italian Ministry of Health, RC08061819 and RC08061822, and by 5X1000 grant 08061821from the San Matteo Hospital. The Microbiology and Virology Unit, *Fondazione IRCCS Policlinico San Matteo*, Pavia, Italy, is supported by grants from the Ministry of Health, RC80206, and by the European Union under grant agreement 101191725 (InFlaMe project, HORIZON-HLTH-2024-DISEASE-08-20 program). A. Ferrari, at the Microbiology and Virology Unit, *Fondazione IRCCS Policlinico San Matteo*, Pavia, Italy, is affiliated to, and supported by, the National PhD Programme in One Health — Approaches to Infectious Diseases and Life Science Research, at the Department of Public Health, Experimental and Forensic Medicine, University of Pavia, Pavia, Italy, and by grant from the Italian Ministry of Health RC8048424. The Cell Factory (P. Comoli, M.A. Avanzini and S. Croce), at *Fondazione IRCCS Policlinico San Matteo*, Pavia, Italy, is supported by grant from the Italian Ministry of Health RC8053724. The Department of Molecular Medicine, University of Padua, Italy, is supported by the European Union’s Horizon 2020 research and innovation program (VEO, grant agreement 874735) and the EU4Health program (DURABLE, grant agreement101102733). The Department of Clinical, Surgical, Diagnostic and Pediatric Sciences, University of Pavia, Italy, and the Department of Molecular Medicine, University of Padua, Italy, are supported by EU funding within the Next-Generation EU-MUR PNRR Extended Partnership initiative on Emerging Infectious Diseases (project no. PE00000007, INF-ACT). The Laboratory of Virology and Infectious Disease (M.R. MacDonald and C.M. Rice) is supported by NIH grants R01AI124690 and R01AI091707. The Murray laboratory is supported by NIH grants R01AI091816, U19AI089992.

## Declaration of interest

J.L.-C. is an inventor on patent application PCT/US2021/042741, filed July 22, 2021, submitted by The Rockefeller University and covering the diagnosis of susceptibility to, and the treatment of, viral disease, and viral vaccines, including COVID-19 and vaccine-associated diseases.

## Author contributions

A.G., F.T., A.F., F.R. S.C., I.C., M.M., J.L.U., E.B., M.An., L.Bi., D.L., C.V., V.C., M.Z., P.C., M.A.A., J.F., M.Au., A.Pi., A.C., S.-Y.Z., J.-L.C. and A.B. performed or supervised experiments, generated and analyzed data, and contributed to the manuscript by providing figures and tables. A.Ma., L.A. and A.C. performed computational analyses of data. F.C., L.Ba., V.G., T.L., D.C., A.A., E.J., P.B., M.R.M., C.M.R., A.Pu., G.R., D.M., Y.S., A.N., D.H., K.O.M., F.B. and J.A. evaluated and recruited WNV patients and controls and/or control cohorts of patients. A.C., S.-Y.Z., J.-L.C. and A.B. supervised the project. J.-L.C. and A.B. conceptualized the project and wrote the manuscript. All the authors edited the manuscript.

## Notes

### Author Declarations

Ethics Committee of San Matteo Research Hospital in Pavia, Italy, EU (Comitato Etico Territoriale Lombardia 6 - Policlinico San Matteo) Ethics Committee of The University Hospital of Padua, Italy, EU Ethics Committee of The University Hospital of Bologna, Italy, EU Ethics Committee of Amedeo di Savoia Hospital-ASL Citta di Torino in Turin, Italy, EU Ethics Committee of the Tel-Aviv Sourasky Medical Center in Israel Ethics Committee of the National Public Health Center in Budapest Ethics Committee of the Rockefeller University in New York, USA The Center for Virology at the Medical University of Vienna, Austria, serves as the national reference center for arboviruses, and has the legal permission to process anonymized patient information on notifiable viral diseases

