## Supplemental figures S1 to S5 for "Autoantibodies neutralizing type I IFNs in 40% of patients with WNV encephalitis in seven new cohorts"

### Slide 1
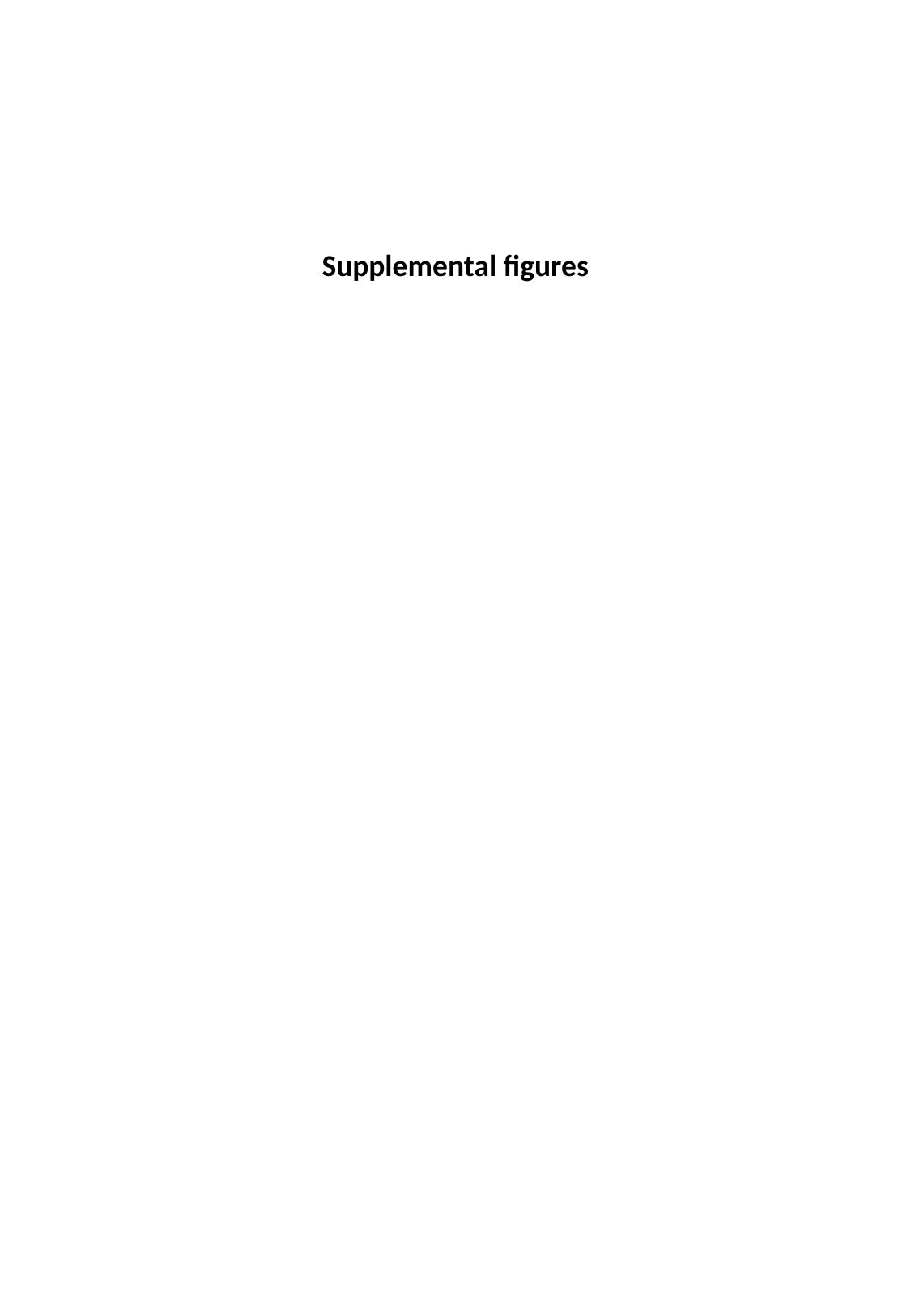

Supplemental figures

### Slide 2
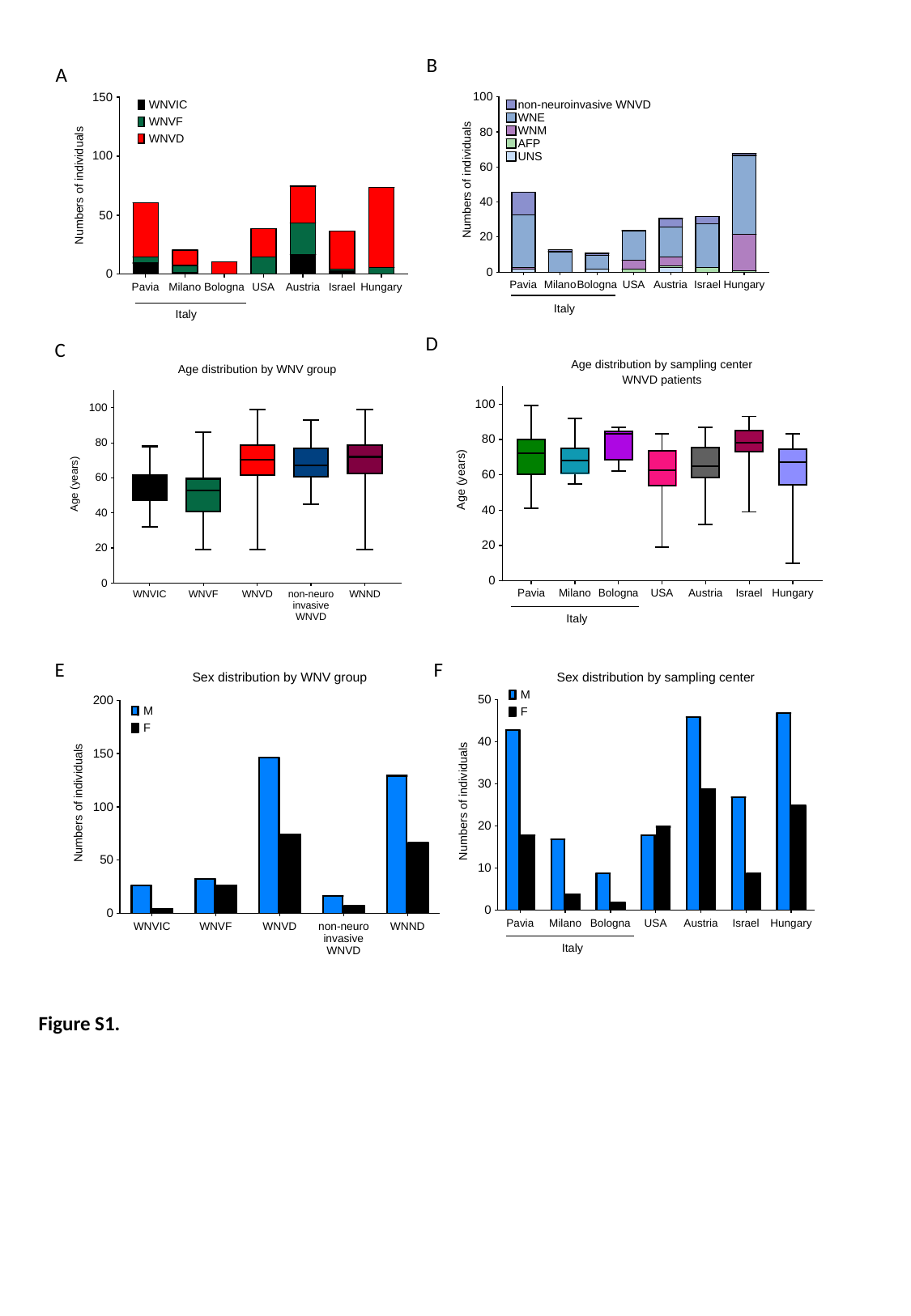

B
A
D
C
E
F
Figure S1.

### Slide 3
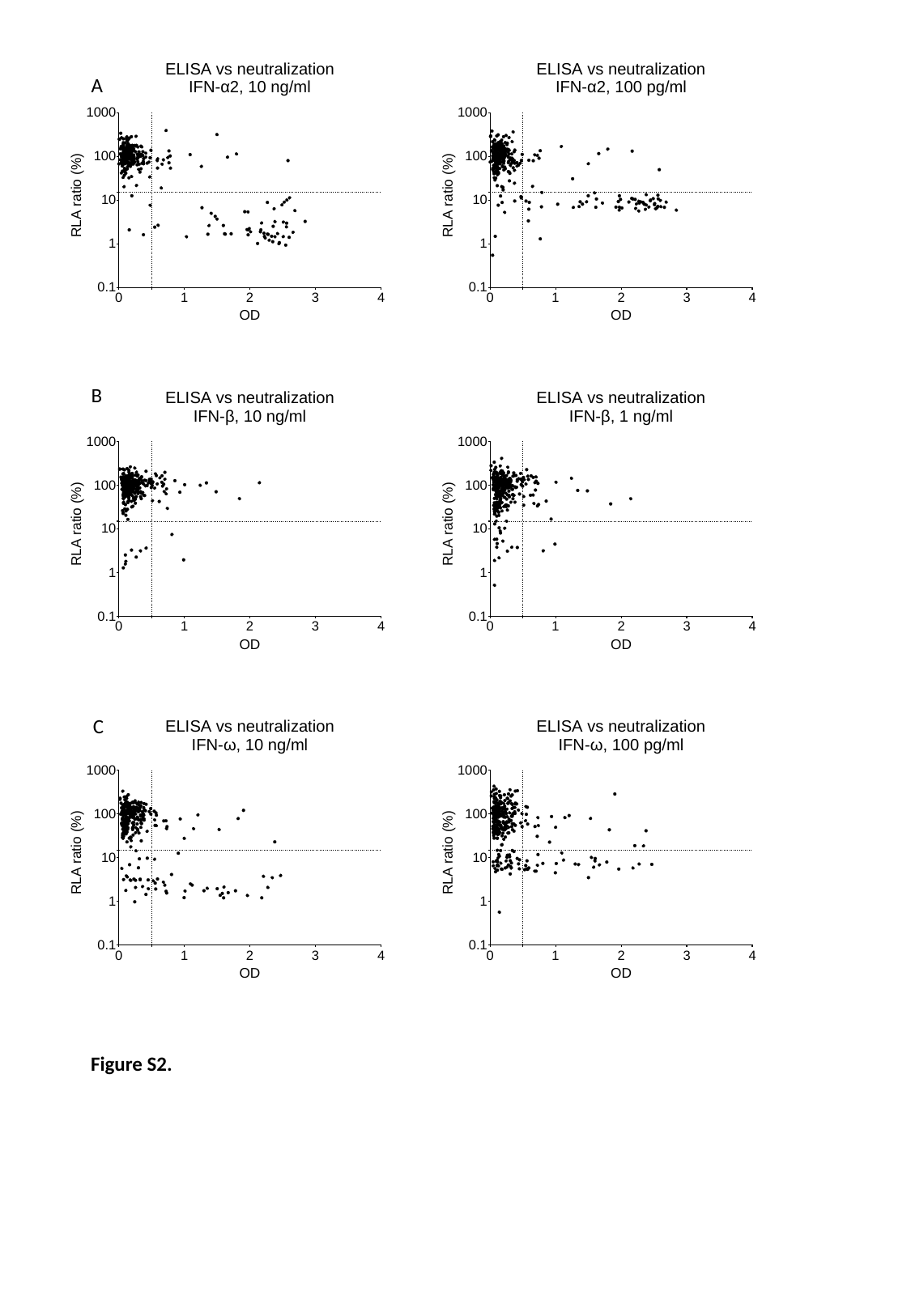

A
B
C
Figure S2.

### Slide 4
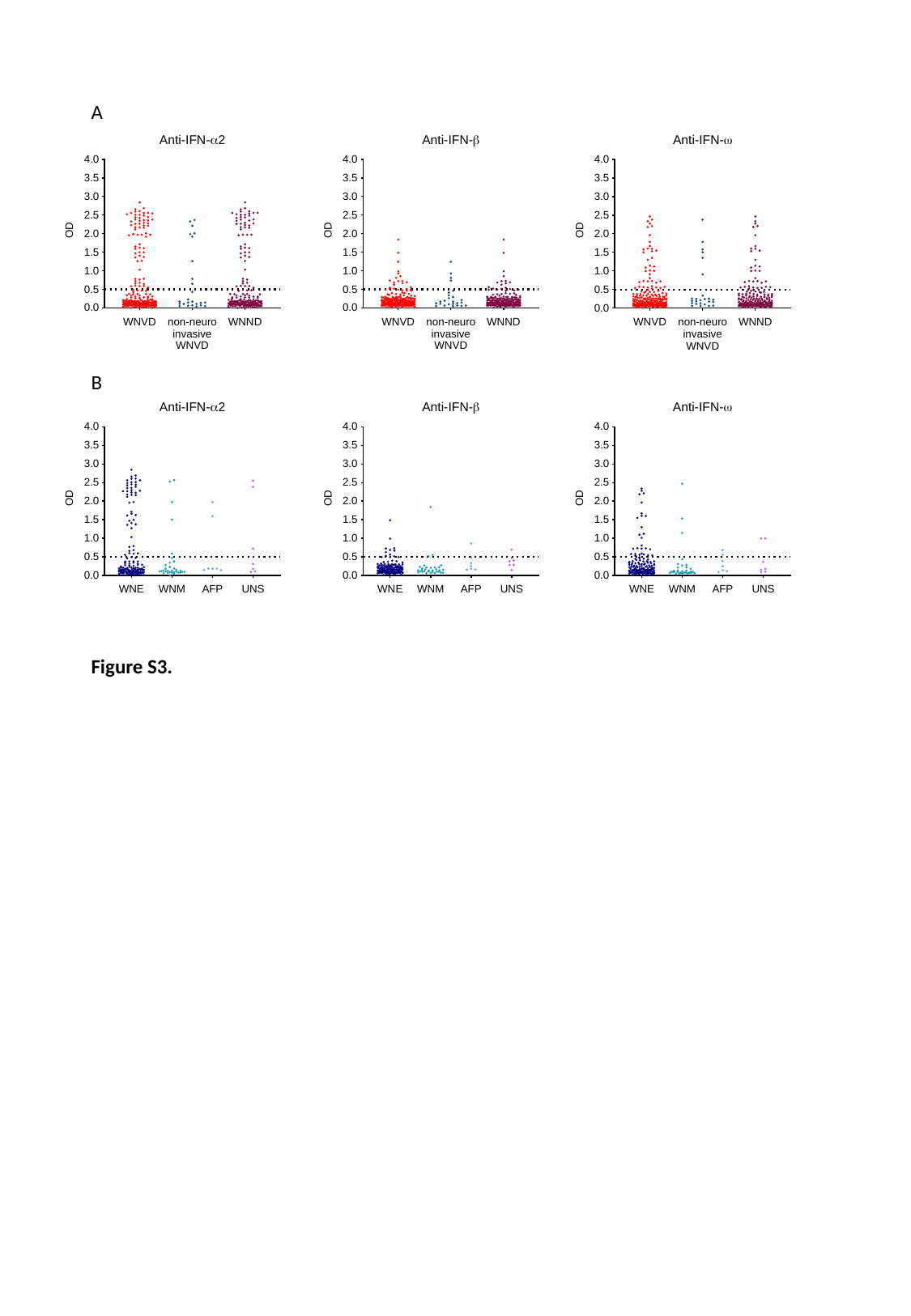

A
B
Figure S3.

### Slide 5
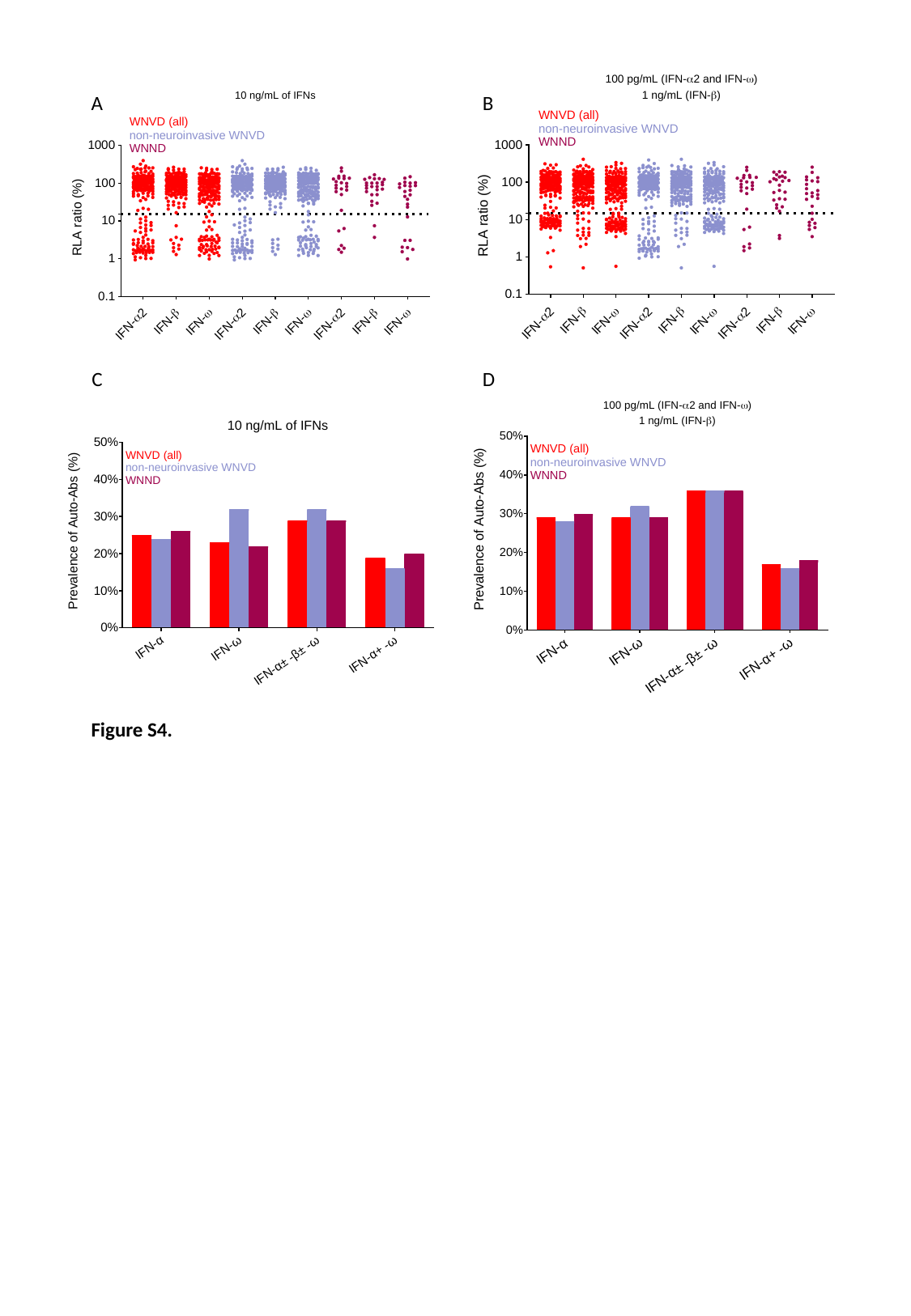

A
B
C
D
Figure S4.

### Slide 6
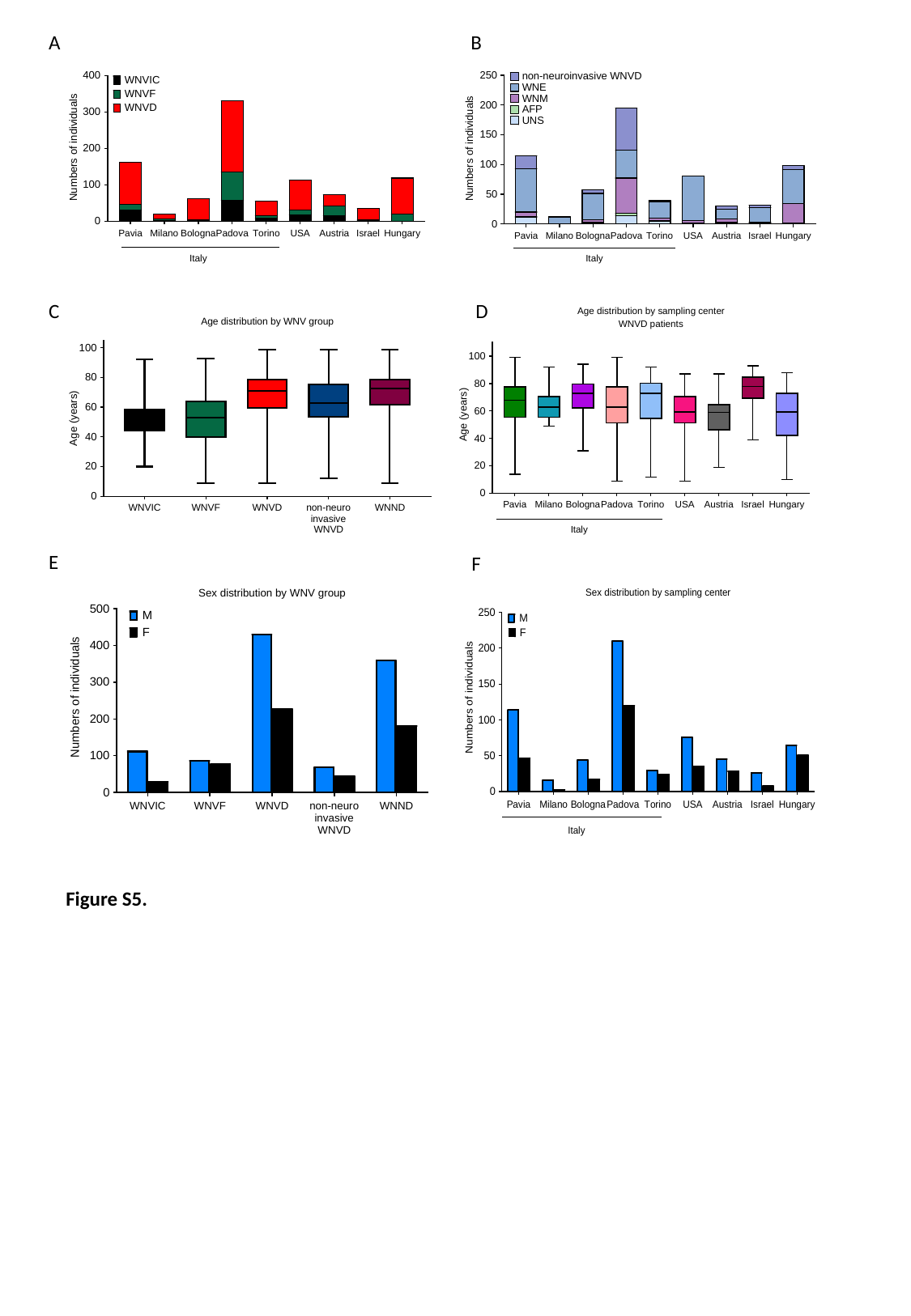

A
B
C
D
E
F
Figure S5.
