## Supplemental tables S1 to S3 for "Autoantibodies neutralizing type I IFNs in 40% of patients with WNV encephalitis in seven new cohorts"

**Supplemental table 1 (table S1). Prevalence of WNVIC, WNVF, WNVD, or WNND of subjects carrying auto-Abs neutralizing at least 1 IFN-I by WNV lineage (WNV-1 or WNV-2).**

| **WNV group** | **WNV-1** | | **WNV-2** | | |
| --- | --- | --- | --- | --- | --- |
|  | **Detected Cases** | **Anti-IFN-α2 (100 pg/ml) and/or anti-IFN-ω (100 pg/ml) and/or anti-IFN-β (1 ng/ml)** | **Detected Cases** | **Anti-IFN-α2 (100 pg/ml) and/or anti-IFN-ω (100 pg/ml) and/or anti-IFN-β (1 ng/ml)** | **P value** |
| WNVIC | 16/66 (24%) | 0/16 (0%) | 50/66 (76%) | 1/50 (2%) |  |
| WNVF | 21/84 (25%) | 1/21 (5%) | 63/84 (75%) | 6/63 (9%) |  |
| WNVD | 69/230 (30%) | 23/69 (33%) | 161/230 (70%) | 58/161 (36%) |  |
| WNND | 56/160 (35%) | 19/56 (34%) | 104/160 (65%) | 48/104 (46%) | 0.81 |
| WNE | 35/56 (63%) | 4/19 (21%) | 53/104 (51%) | 29/53 (55%) | 0.02 |
| WNM | 19/56 (34%) | 14/35 (40%) | 31/104 (30%) | 13/31 (42%) |  |
| AFP | 2/56 (4%) | 1/2 (50%) | 1/104 (1%) | 0/1 (0%) |  |
| UNS | 0/56 (0%) | — | 19/104 (18%) | 6/19 (32%) |  |

WNVIC: West Nile virus infected controls; WNVF: West Nile virus fever; WNVD: West Nile virus disease; WNND: West Nile virus neurological disease; WNE: WNV encephalitis; WNM: WNV meningitis; AFP: acute flaccid paralysis; UNS: unspecified neurological syndrome

Counts or frequency (%)

**Supplemental table 2 (table S2). Risk of WNVIC, WNVF, WNVD, or WNND of subjects carrying auto-Abs neutralizing at least 1 IFN-I by sex.**

| **WNV group** | | **New Cohort** | | | | **New + Old Cohort** | | | |
| --- | --- | --- | --- | --- | --- | --- | --- | --- | --- |
|  |  | **Males** | **Females** | **OR [95%CI]** | **P value** | **Males** | **Females** | **OR [95%CI]** | **P value** |
| WNVIC |  | 1/27 [4%] | 0/5 [0%] | — | 1.0 | 3/114 [3%] | 0/32 [0%] | — | 1.0 |
| WNVF |  | 1/33 [3%] | 0/27 [0%] | — | 1.0 | 11/88 [13%] | 5/80 [6%] | — | 0.17 |
| WNVD |  | 60/147 [41%] | 19/76 [25%] | 2.07 [1.13– 3.89] | 0.02 | 172/433 [40%] | 63/231 [27%] | 1.75 [1.24– 2.48] | 1.7×10⁻³ |
| WNND | | 53/130 [41%] | 17/68 [25%] | 2.06 [1.09– 4.04] | 0.03 | 149/362 [41%] | 56/184 [30%] | 1.59 [1.09– 2.33] | 0.02 |

WNVIC: West Nile virus infected controls; WNVF: West Nile virus fever; WNVD: West Nile virus disease; WNND: West Nile virus neurological disease; WNE: WNV encephalitis; WNM: WNV meningitis; AFP: acute flaccid paralysis; UNS: unspecified neurological syndrome

Counts or frequency (%)

**Supplemental table 3 (table S3). Risk of neuroinvasive disease a and encephalitis for subjects carrying auto-Abs neutralizing specific sets of type I IFNs, relative to the general population, with adjustment for age and sex and risk by age group.**

| **Anti-type I IFN auto-Ab (amount of type I IFN neutralized, in plasma diluted 1:10)** |  | **New cohort** | | **Overall cohort** | |
| --- | --- | --- | --- | --- | --- |
|  | **WNV group** | **OR [95%CI]** | **P value** | **OR [95%CI]** | **P value** |
| Anti-IFN-ω (100 pg/ml) | WNND (All) | 21.7 [15.4-30.6] | < 10^-16^ | 25 [19.8-31.7] | < 10^-16^ |
|  | WNND ≤ 65 | 24.3 [12-49] | < 10^-16^ | 27.4 [17.1-43.7] | < 10^-16^ |
|  | WNND > 65 | 29 [18.9-44.6] | < 10^-16^ | 28.8 [21.6-38.5] | < 10^-16^ |
| Anti-IFN-α2 (100 pg/ml) | WNND (All) | 25 [17.4-35.8] | < 10^-16^ | 29.4 [23-37.6] | < 10^-16^ |
|  | WNND ≤ 65 | 86.6 [41.4-181.2] | < 10^-16^ | 80.1 [44.7-143.6] | < 10^-16^ |
|  | WNND > 65 | 22.5 [14.7-34.5] | < 10^-16^ | 26.2 [19.8-34.7] | < 10^-16^ |
| Anti-IFN-α2 (100 pg/ml) and/or anti-IFN-ω (100 pg/ml) and/or anti-IFN-β (10 ng/ml) | WNND (All) | 16.8 [12.1-23.4] | < 10^-16^ | 18.6 [14.9-23.2] | < 10^-16^ |
|  | WNND ≤ 65 | 24.4 [13.7-43.7] | < 10^-16^ | 20.7 [13.7-31.3] | < 10^-16^ |
|  | WNND > 65 | 18.1 [12-27.3] | < 10^-16^ | 20.2 [15.4-26.6] | < 10^-16^ |
| Anti-IFN-α2 (100 pg/ml) and anti-IFN-ω (100 pg/ml) | WNND (All) | 42.2 [27.8-63.9] | < 10^-16^ | 53.6 [39.4-72.9] | < 10^-16^ |
|  | WNND ≤ 65 | 378.8 [96.3-1490] | 8.9 x 10^-16^ | 383.1 [131-1120.4] | < 10^-16^ |
|  | WNND > 65 | 40.6 [25.4-64.7] | < 10^-16^ | 42.9 [30.8-59.7] | < 10^-16^ |
| Anti-IFN-ω (10 ng/ml) | WNND (All) | 48.7 [32.5-73.1] | < 10^-16^ | 56.8 [42.7-75.6] | < 10^-16^ |
|  | WNND ≤ 65 | 100.6 [40.2-251.6] | 2.6 x 10^-11^ | 134.7 [74.5-243.6] | < 10^-16^ |
|  | WNND > 65 | 45.8 [29.3-71.7] | 9.2 x 10^-12^ | 44.5 [32.4-60.9] | < 10^-16^ |
| Anti-IFN-α2 (10 ng/ml) | WNND (All) | 51.6 [34.8-76.4] | < 10^-16^ | 65.9 [50.2-86.5] | < 10^-16^ |
|  | WNND ≤ 65 | 273.4 [119.9-623.0] | < 10^-16^ | 245.3 [129.9-463.4] | < 10^-16^ |
|  | WNND > 65 | 39.5 [25.4-61.4] | < 10^-16^ | 49.4 [36.8-66.4] | < 10^-16^ |
| Anti-IFN-α2 (10 ng/ml) and/or anti-IFN-ω (10 ng/ml) and/or anti-IFN-β (10 ng/ml) | WNND (All) | 28 [19.1-41] | < 10^-16^ | 32.8 [24.9-43.3] | < 10^-16^ |
|  | WNND ≤ 65 | 46.4 [21.6-99.6] | 2.2 x 10^-14^ | 45.7 [26.1-79.9] | < 10^-16^ |
|  | WNND > 65 | 27.7 [17.7-43.4] | < 10^-16^ | 31.1 [22.5-42.9] | < 10^-16^ |
| Anti-IFN-α2 (10 ng/ml) and anti-IFN-ω (10 ng/ml) | WNND (All) | 101.3 [63.3-162.1] | < 10^-16^ | 123.5 [85.6-178.3] | < 10^-16^ |
|  | WNND ≤ 65 | 442 [128.6-1518.6] | 2.5 x 10^-14^ | 602 [224.2-1616.2] | < 10^-16^ |
|  | WNND > 65 | 82.9 [50.2-137] | < 10^-16^ | 84.6 [57.6-124.3] | < 10^-16^ |
| Anti-IFN-α2 (10 ng/ml) and anti-IFN-ω (100 pg/ml) | WNND (All) | 59.0 [34.8-100.2] | < 10^-16^ | 83.1 [53.8-128.5] | < 10^-16^ |
|  | WNND ≤ 65 | 1616.4 [84.1-31074.1] | 2.3 x 10^-13^ | 1841.1 [109.5-30940.6] | < 10^-16^ |
|  | WNND > 65 | 48.6 [27.9-84.8] | < 10^-16^ | 58.4 [37.5-90.9] | < 10^-16^ |
| Anti-IFN-α2 (10 ng/ml) and anti-IFN-ω (10 ng/ml) and anti-IFN-β (10 ng/ml) | WNND (All) | 243.9 [12.5-4744.1] | 2.9 x 10^-6^ | 142.0 [7.9-2535.1] | 1.1 x 10^-7^ |
|  | WNND ≤ 65 | 113.1 [4.5-2826.3] | 3 x 10^-3^ | 46.3 [1.9-1146.1] | 1.0 x 10^-2^ |
|  | WNND > 65 | 159.5 [7.5-3412.3] | 2 x 10^-4^ | 110.4 [6-2020.6] | 2.2 x 10^-6^ |
| Anti-IFN-ω (100 pg/ml) | WNE (All) | 23.7 [16.3-34.6] | < 10^-16^ | 22.8 [17.4-29.8] | < 10^-16^ |
|  | WNE ≤ 65 | 29.7 [13.1-67.3] | 9.2 x 10^-10^ | 27.9 [15.8-49.1] | < 10^-16^ |
|  | WNE > 65 | 29.5 [18.8-46.4] | < 10^-16^ | 25.6 [18.6-35.4] | < 10^-16^ |
| Anti-IFN-α2 (100 pg/ml) | WNE (All) | 26.1 [17.6-38.7] | < 10^-16^ | 25.1 [19.0-33.2] | < 10^-16^ |
|  | WNE ≤ 65 | 93.5 [40.5-215.9] | < 10^-16^ | 73.2 [37.5-143.1] | < 10^-16^ |
|  | WNE > 65 | 23.3 [14.9-36.5] | < 10^-16^ | 22.5 [16.4-30.8] | < 10^-16^ |
| Anti-IFN-α2 (100 pg/ml) and/or anti-IFN-ω (100 pg/ml) and/or anti-IFN-β (10 ng/ml) | WNE (All) | 17.4 [12.1-25.1] | < 10^-16^ | 16.4 [12.7-21.2] | < 10^-16^ |
|  | WNE ≤ 65 | 25.6 [12.7-51.3] | 1.1 x 10^-12^ | 19.5 [11.8-32.4] | < 10^-16^ |
|  | WNE > 65 | 18.5 [11.9-28.7] | < 10^-16^ | 17.3 [12.7-23.5] | < 10^-16^ |
| Anti-IFN-α2 (100 pg/ml) and anti-IFN-ω (100 pg/ml) | WNE (All) | 46.4 [29.8-72.2] | < 10^-16^ | 46.4 [33.1-65.0] | < 10^-16^ |
|  | WNE ≤ 65 | 705.6 [138.6-3592.6] | 3.3 x 10^-15^ | 420.7 [128.5-1377.3] | < 10^-16^ |
|  | WNE > 65 | 42.1 [25.8-68.6] | < 10^-16^ | 38.3 [26.6-55] | < 10^-16^ |
| Anti-IFN-ω (10 ng/ml) | WNE (All) | 50.4 [32.6-78.1] | < 10^-16^ | 48.6 [35.2-67.2] | < 10^-16^ |
|  | WNE ≤ 65 | 142.9 [51.7-394.8] | 1.5 x 10^-10^ | 142.8 [70.1-291] | < 10^-16^ |
|  | WNE > 65 | 45.8 [28.5-73.5] | < 10^-16^ | 39.3 [27.7-55.8] | < 10^-16^ |
| Anti-IFN-α2 (10 ng/ml) | WNE (All) | 52.0 [34.1-79.5] | < 10^-16^ | 55.6 [40.9-75.5] | < 10^-16^ |
|  | WNE ≤ 65 | 304.8 [118.7-782.8] | < 10^-16^ | 268.9 [128.8-561.5] | < 10^-16^ |
|  | WNE > 65 | 41.3 [26-65.6] | < 10^-16^ | 42.5 [30.6-59] | < 10^-16^ |
| Anti-IFN-α2 (10 ng/ml) and/or anti-IFN-ω (10 ng/ml) and/or anti-IFN-β (10 ng/ml) | WNE (All) | 29.1 [19.2-44.0] | < 10^-16^ | 28.2 [20.6-38.4] | < 10^-16^ |
|  | WNE ≤ 65 | 54.5 [22.6-131.6] | 1.3 x 10^-11^ | 44.3 [22.8-86.2] | < 10^-16^ |
|  | WNE > 65 | 27.8 [17.4-44.6] | < 10^-16^ | 26.4 [18.6-37.6] | < 10^-16^ |
| Anti-IFN-α2 (10 ng/ml) and anti-IFN-ω (10 ng/ml) | WNE (All) | 106.7 [65.0-175.0] | < 10^-16^ | 105.8 [71.2-157.3] | < 10^-16^ |
|  | WNE ≤ 65 | 554.5 [145.9-2107.5] | 6.8 x 10^-13^ | 649.6 [217.9-1937] | < 10^-16^ |
|  | WNE > 65 | 87 [51.6-146.8] | < 10^-16^ | 76.1 [50.3-115.1] | < 10^-16^ |
| Anti-IFN-α2 (10 ng/ml) and anti-IFN-ω (100 pg/ml) | WNE (All) | 65.9 [38.1-114.2] | < 10^-16^ | 71.8 [45.3-113.7] | < 10^-16^ |
|  | WNE ≤ 65 | 2597.5 [117.1-57597.3] | 3.9 x 10^-12^ | 2218.4 [125.1-39337.7] | < 10^-16^ |
|  | WNE > 65 | 54 [30.4-95.8] | < 10^-16^ | 52.1 [32.6-83.3] | < 10^-16^ |
| Anti-IFN-α2 (10 ng/ml) and anti-IFN-ω (10 ng/ml) and anti-IFN-β (10 ng/ml) | WNE (All) | 218.5 [10.3-4629.5] | 6.0 x 10^-5^ | 128.0 [6.8-2414.0] | 5.1 x 10^-6^ |
|  | WNE ≤ 65 | 147.6 [5.9-3717.3] | 2 x 10^-3^ | 63.2 [2.5-1569] | 8 x 10^-3^ |
|  | WNE > 65 | 80.7 [3.3-2004.6] | 5 x 10^-3^ | 74.8 [3.8-1455.8] | 1.6 x 10^-4^ |

WNND: West Nile neuroinvasive disease, a subgroup of WNVD; WNE: West Nile Encephalitis, a subgroup of WNND. ≤65 and >65 indicate age cut-offs; anti-IFN-ω and anti-IFN-α2 indicate auto-Abs neutralizing IFN-ω or IFN-α2, respectively, regardless of their effects on other IFNs
